# Rapid Identification and Phenotyping of Nonalcoholic Fatty Liver Disease Patients Using an Algorithmic Approach in Diverse, Urban Healthcare Systems

**DOI:** 10.1101/2021.04.27.21256139

**Authors:** Anna O. Basile, Anurag Verma, Leigh Anne Tang, Marina Serper, Andrew Scanga, Ava Farrell, Brittney Destin, Rotonya M. Carr, Anuli Anyanwu-Ofili, Gunaretnam Rajagopal, Abraham Krikhely, Marc Bessler, Muredach P. Reilly, Marylyn D. Ritchie, Nicholas P. Tatonetti, Julia Wattacheril

## Abstract

**Objectives:** Nonalcoholic Fatty Liver Disease (NAFLD) is the most common global cause of chronic liver disease. Therapeutic interventions are rapidly advancing for its inflammatory phenotype, nonalcoholic steatohepatitis (NASH) at all stages of disease. Diagnosis codes alone fail to accurately recognize and stratify at-risk patients. Our work aims to rapidly identify NAFLD patients within large electronic health record (EHR) databases for automated stratification and targeted intervention based on clinically relevant phenotypes.

**Methods:** We present a rule-based phenotyping algorithm for the rapid identification of NAFLD patients developed using EHRs from 6.4 million patients at Columbia University Irving Medical Center (CUIMC) and validated at two independent healthcare centers. The algorithm uses the Observational Medical Outcomes Partnership (OMOP) Common Data Model and queries multiple structured and unstructured data elements, including diagnosis codes, laboratory measurements, radiology and pathology modalities.

**Results:** Our approach identified 16,006 CUIMC NAFLD patients, 10,753 (67%) of whom were previously unidentifiable by NAFLD diagnosis codes. Fibrosis scoring on patients without histology identified 943 subjects with scores indicative of advanced fibrosis (FIB-4, APRI, NAFLD–FS). The algorithm was validated at two independent healthcare systems, University of Pennsylvania Health System (UPHS) and Vanderbilt Medical Center (VUMC), where 20,779 and 19,575 NAFLD patients were identified, respectively. Clinical chart review identified a high positive predictive value (PPV) across all healthcare systems: 91% at CUIMC, 75% at UPHS, and 85% at VUMC, and a sensitivity of 79.6%.

**Conclusions:** Our rule-based algorithm provides an accurate, automated approach for rapidly identifying, stratifying, and sub-phenotyping NAFLD patients within a large EHR system.

**Study Highlights:** *WHAT IS KNOWN:*

- NAFLD is the leading form of chronic liver disease with a rising prevalence in the population.
- NAFLD is often under-recognized in at-risk individuals, including within healthcare settings.
- Current means of identification and stratification are complex and dependent on provider recognition of clinical risk factors.

*WHAT IS NEW HERE:*

- An accurate, validated rule-based algorithm for the high-throughput and rapid EHR identification of NAFLD patients.
- Rapid discovery of a NAFLD cohort from diverse EHR systems comprising approximately 12.1 million patients.
- Our algorithm has high performance (mean PPV=85%, sensitivity=79.6%) in NAFLD patient discovery.
- The majority of algorithmically derived NAFLD patients were previously unidentified within healthcare systems.
- Computational stratification of individuals with advanced fibrosis can be achieved rapidly.

## Introduction

NAFLD is the most common form of chronic liver disease worldwide affecting 25-30% of the general adult population in industrialized countries (^1^). NAFLD, along with its inflammatory NASH, is often under-diagnosed (^2^) due to the cost and invasiveness of liver biopsy, the current gold standard of diagnosis. Identifying and stratifying NAFLD patients is critically important to effective healthcare delivery from diabetes prevention and treatment to targeted diagnostics, specialist referral, cancer screening, genomic analyses and intervention for longitudinal assessment and follow-up. Early recognition is particularly important given disease model projections of a doubling or tripling of end-stage liver disease patients by 2030 in many parts of the world (^3^). Prioritizing preventative care for groups at high risk of progression, such as those exhibiting NASH and advanced fibrosis, is crucial given the association with clinical outcomes (^4^).

Emerging therapies for NASH will be limited in application if at-risk individuals remain difficult to identify in health care systems. Unfortunately, given current limitations inherent in diagnostic coding for this disease, the rapid identification of patients with NAFLD is problematic. Advances in circulating blood biomarkers and imaging biomarkers can assist in risk stratifying patients with identified NAFLD, particularly those with advanced fibrosis. Electronic health record (EHR) phenotyping is another means by which patients can be targeted for diagnosis and risk stratification. EHR records are collected prospectively in a large-scale, long-term follow-up manner (^5^) and can provide the data needed to phenotypically identify patients. These properties, along with the inclusion of diverse aspects of patients’ health-related information make EHRs a valuable data source for phenotype discovery. EHRs are limited by the completeness and accuracy of data which may have confounding effects if not properly addressed in the study design (^6–8^). One approach in addressing these inaccuracies is to use a wide range of different data sources available in the EHR (including structured data, such as diagnosis/billing codes and laboratory measures, as well as unstructured elements such as imaging/radiology reports and provider notes) as a means of diagnostic confirmation. Additionally, quality control parameters can be implemented to reduce false-positive identifications.

Herein, we describe a rule-based phenotype algorithm developed at Columbia University Irving Medical Center (CUIMC) which expands on earlier work (^9–12^) by utilizing a multitude of EHR data sources (structured and unstructured) to identify NAFLD and NASH patients for clinical intervention. The algorithm queries over 400 diagnosis codes, 100 laboratory and serology measurements, pathology, and various radiology modalities. To demonstrate cross-institutional utility and performance, we validate the algorithm at two large independent medical centers. We also perform fibrosis scoring tests on all CUIMC identified NAFLD patients without histologically-confirmed NASH, identifying patients at highest risk for progressing to end-stage liver outcomes and demonstrating the feasibility of rapid risk stratification techniques. As this algorithm was developed using the Observational Medical Outcomes Partnership (OMOP) Common Data Model (CDM), it can easily be deployed at healthcare institutions that support the OMOP CDM, which is presently over 90 sites worldwide, including the National Institutes of Health *All of Us* Research Program (^13^).

## Methods

Our NAFLD algorithm was developed using EHR data within the CUIMC health care center. CUIMC serves the diverse population of New York City, and is composed of approximately 38% Hispanic patients, 37% European American, 21% African American, and 4% other ethnicities. Observational Health Data Sciences and Informatics (OHDSI) is an international initiative with over 3,000 collaborators focused on improving the use of healthcare data (^14,15^). OHDSI maintains the Observational Medical Outcomes Partnership (OMOP) Common Data Model (CDM) (^16^), a data standard that normalizes observational data to enable efficient, reproducible analyses. CUIMC is a participant of OHDSI and maintains an OMOP database of primary and ancillary EHR systems, pharmacy, and billing systems. The CUIMC clinical data warehouse (CDW) encompasses this OMOP database and other clinical data distributed for research purposes, such as provider notes, unstructured, and imaging data (^17^). At the time of this study, the CDW contained records for 6.4 million patients dating back to 1985. Figure 1 provides an illustrative depiction of the NAFLD algorithm development and validation process.

**Figure 1:**
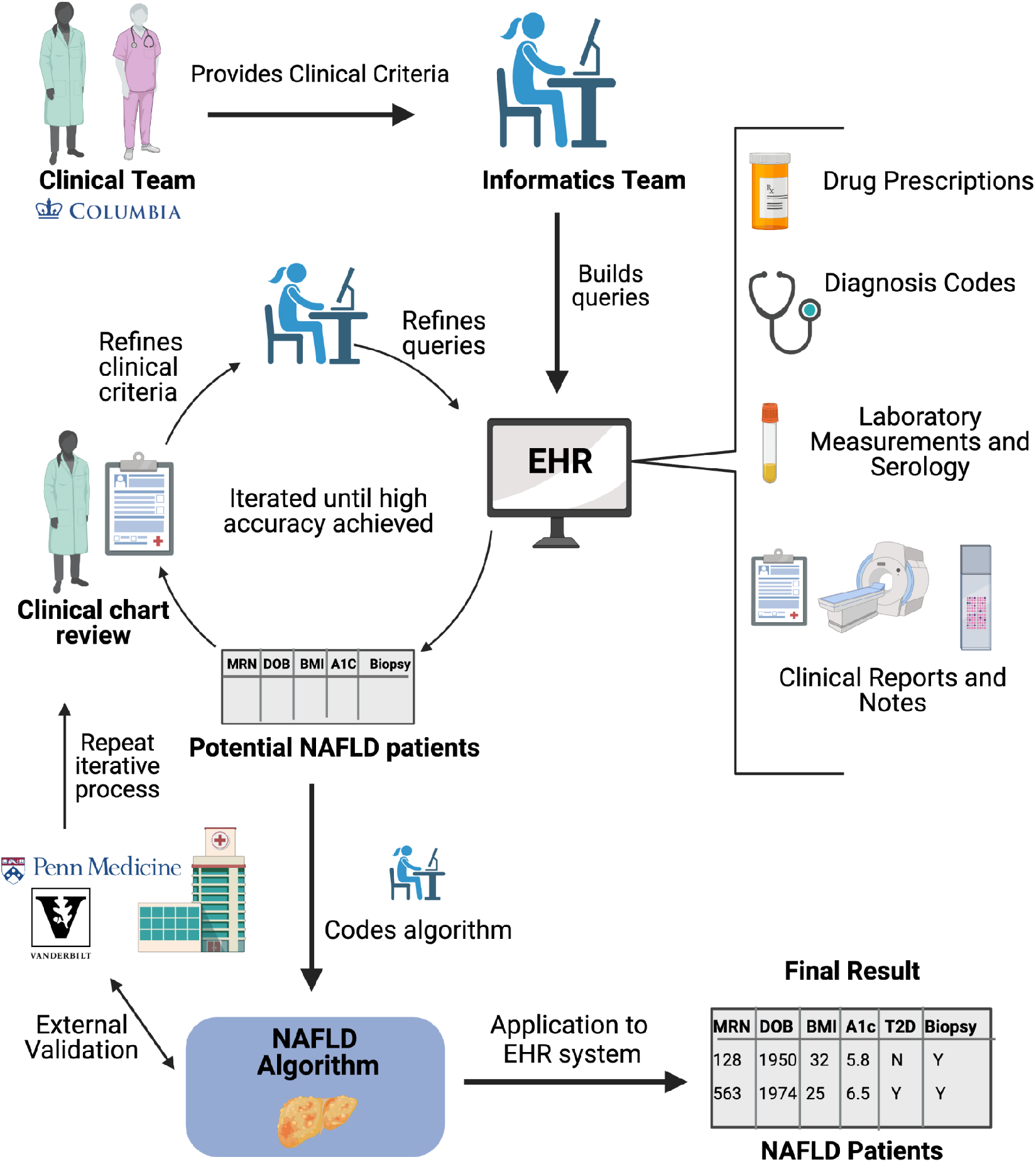
Illustration of the NAFLD algorithm development and validation process. The algorithm was developed at Columbia University Irving Medical Center (CUIMC) by clinical and informatics teams. Clinical criteria for the algorithm, provided by medical experts, was used by the bioinformatics team to design queries which produced a set of potential NAFLD patients. The charts of these patients were reviewed by the clinical team to determine true NAFLD status. Clinical criteria, as represented in the EHR system, was adjusted based on chart review results and the queries were refined. This process was repeated across each step of the algorithm until high accuracy was achieved. Once achieved, the queries were used to code the algorithm. Algorithmic validation was performed at the University of Pennsylvania Health System (UPHS) and Vanderbilt Medical Center (VUMC) where the iterative process described above was repeated by clinical and informatic experts at each site. The final output of the algorithm is a list of NAFLD patients along with clinical characteristics (subset depicted above). EHR = Electronic Health Record; A1c=Glycated hemoglobin; T2D=Type 2 Diabetes; Y= Yes; N= No; DOB=Date of Birth. Stock images for this figure are from BioRender.com.

Code for the NAFLD algorithm predominantly consists of SQL queries of the structured OMOP database coupled with unstructured data parsing of notes from pathology and radiology reports. The algorithm has three main steps which each flow consecutively (Figure 2): 1). Inclusion of potential NAFLD patients, 2). Removal of non-NAFLD patients meeting stringent exclusion criteria, 3). Verification of hepatic steatosis. Please see the Supplement for extended methods.

**Figure 2:**
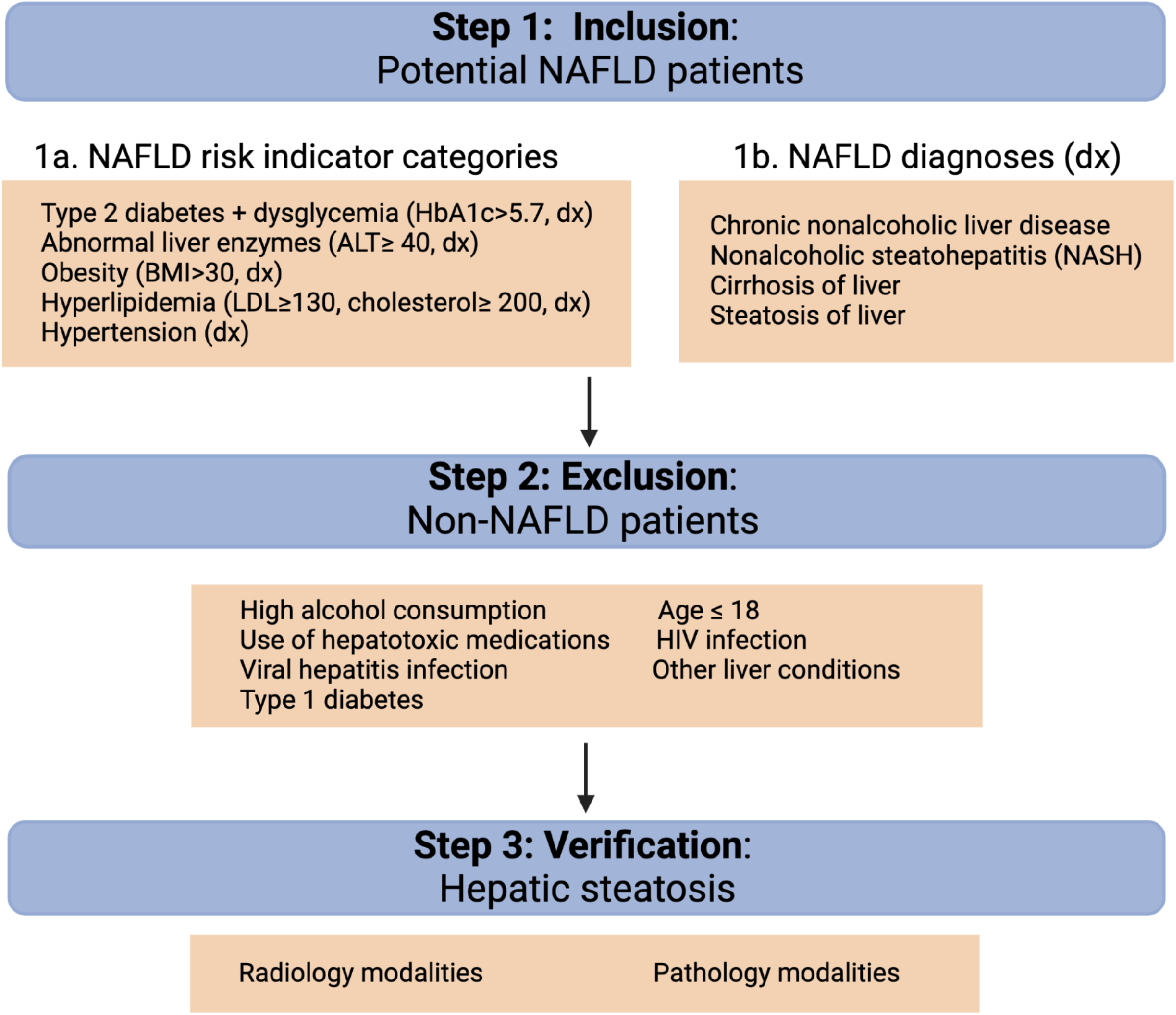
Three main steps of the NAFLD algorithm. Step 1a lists the NAFLD risk indicator categories that were used for patient identification and lists a few examples of selection criteria. A complete list of codes for selection or exclusion criteria can be found in the following Supplementary Tables: Step 1a (S Table 1), Step 1b (S Table 2), Step 2 (S Table 3), and Step 3 (S Table 4 and S Table 5). dx = diagnosis code (International Classification of Diseases, Ninth and Tenth Revision (ICD9/10)).

### Step 1: Identification of NAFLD patients

Step 1 identifies NAFLD patients by the presence of a NAFLD risk indicator and/or a NAFLD diagnosis (see Supplementary Tables 1 and 2 (S1, S2) for codes). NAFLD risk indicators include: type 2 diabetes and dysglycemia (Table S1a), obesity (Table S1b), abnormal liver enzymes (Table S1c), hyperlipidemia (Table S1d), or hypertension (Table S1e).

### Step 2: Exclusion of patients with confounding diagnoses

Cases meeting specified exclusion criteria (see Supplementary Table 3 (S3)) were removed in Step 2. Exclusion criteria include excessive alcohol use, confounding liver or extrahepatic conditions that may result in secondary hepatic steatosis, including viral hepatitis, type 1 diabetes, and others. Detailed exclusion as often undertaken for clinical trials enrollment was codified. Patients prescribed a hepatotoxic medication associated with steatosis (^18^), such as anti-retrovirals, were also excluded.

### Step 3: Verification of hepatic steatosis

Radiology and pathology reports from 1980-2019 were used to verify hepatic steatosis. Regular expressions, a powerful pattern search language and tool (^19^), were used in conjunction with key terms to identify language and usage context indicative of hepatic steatosis in a string-matching approach. Language for an indicator of NAFLD and NASH were included (Supplementary Tables 4 (S4) and 5 (S5)).

### Fibrosis Scoring

To identify patients at risk for fibrotic NAFLD, we applied 3 common fibrosis scoring metrics on patients lacking histology: Fibrosis-4 (FIB-4)(^20^), aspartate transaminase to Platelet Ratio Index (APRI) (^21^), and the NAFLD Fibrosis score (^22^) calculation (Supplementary equations 1-3). Data for these calculations were extracted from patient clinical records at cross-sectional timepoints. To ‘rule in’ advanced fibrosis, we required patients to exhibit an elevated score in at least 2 metrics (FIB-4 > 3.25, an APRI >1.0, and a NAFLD FS > 0.675).

### Quality Control

To minimize EHR diagnosis errors, we employed quality control (QC) measures requiring patients to have ≥2 NAFLD risk indicators, a risk indicator and a NAFLD diagnosis, or ≥3 unique occurrences of a single given NAFLD risk indicator. The cohort was restricted to patients 18 or older (using earliest date of hepatic steatosis confirmation).

### Algorithm Validation

The algorithm was validated at two external, independent healthcare systems, University of Pennsylvania Health System (UPHS) in Philadelphia, Pennsylvania and Vanderbilt Medical Center (VUMC) in Nashville, Tennessee. UPHS maintains a data warehouse for translational research that combines data (discrete and unstructured text reports) from five hospitals in the greater Philadelphia area and one in Princeton, New Jersey. The CDW contains data for 3 million patients dating back to 2005. This population is 64% European American, 24% African American, and 12% other ethnicities. At the time of the study, UPHS used an Epic EHR platform and validation was performed using codes and terminologies found in the Supplement. VUMC maintains a de-identified data warehouse dating to 1990 that contains both structured and unstructured data and describes 3.2 million patients, of which 82% are European American, 13% African American, 4% Hispanic, and 1% other ethnicities. VUMC maintains an OMOP database and validation at this site was performed using data standardized to the OMOP CDM.

### Chart Review and Algorithmic Performance

Manual, retrospective chart review was performed to review data elements for cohort construction and to assess algorithmic accuracy. This clinical review served as a critical component of algorithm development, allowing us to evaluate the efficacy of adding or removing selection criteria across the algorithm. It allowed us to adjust NAFLD risk indicator criteria and keyword terminology indicative of hepatic steatosis. Over 150 MRNs were reviewed at CUIMC during development by two clinical research coordinators and verified by a board-certified transplant hepatologist. Chart review at VUMC and UPHS was similarly performed by board-certified transplant hepatologists.

Chart review was also used in the calculation of algorithmic positive predictive value (PPV) at each of the assessed sites. PPV is the proportion of patients identified by the phenotyping algorithm as having the condition, determined by expert chart review. We reviewed the charts of 200 patients, independent of those assessed for diagnostic code selection to calculate PPV at CUIMC. Hepatologists at VUMC and UPHS reviewed 20 charts for PPV calculation. Overall 390 clinical charts were reviewed during algorithm development and validation.

We performed sensitivity analysis to determine our algorithm’s ability to identify true NAFLD patients using a NAFLD registry maintained by CUIMC hepatology with records from 2006 to 2019. We define sensitivity as the number of patients within this registry who were correctly identified by our algorithm. We assessed sensitivity across three different categories: using all patients within this registry, restricting to patients for whom there was complete data (structured and unstructured) within the CDW, and omitting patients for whom exclusion diagnosis codes could not be physician-verified. Sensitivity analyses were performed solely at CUIMC as we had complete data access within this institution allowing us to thoroughly query medical records and robustly interact with the EHR.

## Results

Our algorithm identified 16,006 NAFLD patients with verified hepatic steatosis at CUIMC with 67% of this cohort previously undiagnosed by NAFLD codes (Figure 3). Over 40% of these patients self-identified as Black, Hispanic, or Asian (Table 1). The algorithm identified 20,779 NAFLD UPHS patients and 19,575 VUMC patients (Figure 3). The majority of patients initially meeting criteria for inclusion were dropped from the algorithm during verification of hepatic steatosis. Of the total potential NAFLD patients, 3.2% at CUIMC, 3.1% at UPHS, and 7.5% of patients at VUMC had algorithmic verified steatosis indicative of NAFLD. This large drop in sample size was primarily due to a lack of available imaging or biopsy data within each system’s CDW. All 16,006 NAFLD patients identified at CUIMC with a NAFLD diagnosis code were also diagnosed with a risk indicator (e.g., abnormal liver enzymes, obesity) (Table 2).

**Figure 3:**
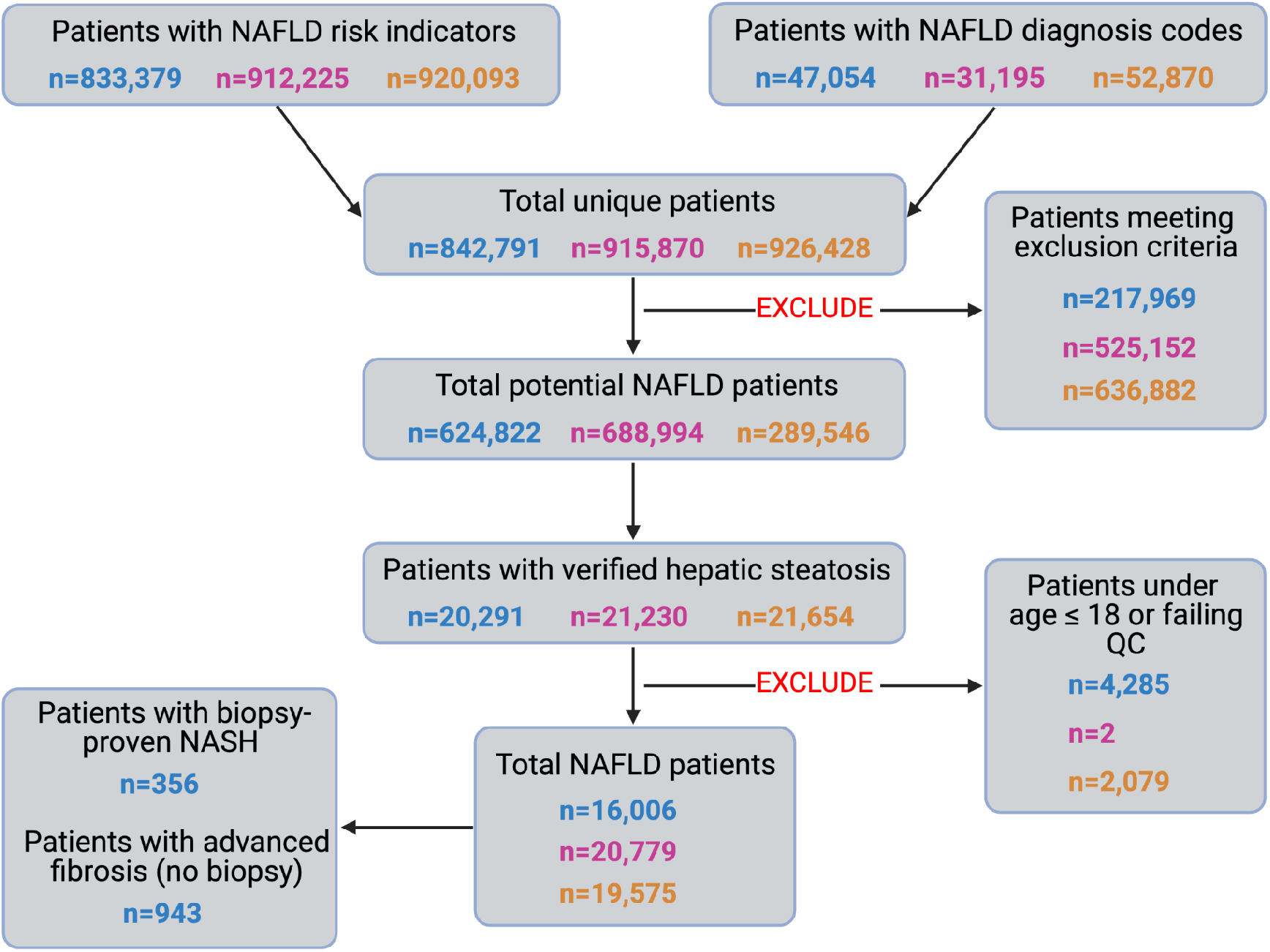
Counts of patients at each stage of the algorithm. Data from Columbia University Irving Medical Center (CUIMC) is in blue, that from University of Pennsylvania Healthcare System (UPHS) is in pink/purple, and numbers from Vanderbilt Medical Center (VUMC) are in orange.

**Table 1:** Demography and summary information for identified NAFLD patients. F =Female; M= male; U=unknown/undeclared. Age of diagnosis is based on the earliest date of verified hepatic steatosis. Mean age is noted with standard deviation in parentheses. Race/ethnicity values may not aggregate to 100% as some sites code race and ethnicity separately.

|  | CUIMC | UPHS | VUMC |
| --- | --- | --- | --- |
| **Mean age at diagnosis**<br>**(+/- Standard Deviation)** | 57.4 (+/- 15.7) | 57.0 (+/- 15.4) | 52.3 (+/- 16.8) |
| **Biological Sex** | F=55.7% M=44.3% | F=44.2%<br>M=55.8% | F=44.4 M=55.6% U=0.01% |
| **Type 2 diabetes** | 53.3% | 31.0% | 24.9% |
| **Obesity** | 45.2% | 60% | 48.8% |
| **Percent Race/Ethnicity** |  |  |  |
| White | 33.9% | 68.4% | 79.3% |
| Black | 9.0% | 16.1% | 12.5% |
| Hispanic | 31.1% | White = 4.8%<br>Black = 1.2% | 3.7% |
| Asian | 1.7% | 2.7% | 0.9% |
| Other | 0.2% | 4.0% | 1.1% |
| Unknown | 24.1% | 2.9% | 6.0% |

**Table 2:** Counts of patients with diagnoses in each risk indicator category of the total identified NAFLD patients (n=16,006) at CUIMC.

| **Risk Indicator Category** | **Count of Patients with Diagnosis** |
| --- | --- |
| Abnormal Liver Enzymes | 5,466 |
| Obesity | 7,271 |
| Type 2 Diabetes and Dysglycemia | 8,658 |
| Hyperlipidemia | 10,690 |
| Hypertension | 11,717 |

Previous studies have shown that using a diagnostic combination of obesity, type 2 diabetes (T2D), and abnormal ALT values predicts NAFLD with high accuracy (^23^). Using a simpler algorithm consisting only of these three diagnoses, we identified 2,107 NAFLD patients at CUIMC, missing 86.9% of the patients discovered by our more complex algorithm. Additionally, 38.8% of these 2,107 patients met at least one exclusion criteria included in our algorithm (e.g., alcohol abuse, viral hepatitis infection). Thus, the combination of obesity, DM2 and abnormal ALT is helpful in identifying patients at risk for NAFLD, but may not necessarily yield true NAFLD patients. The additive value of a phenotyping algorithm of the complexity presented here is in the recognition of patients for whom elevated ALT, obesity, and DM2 was not observed. In diverse populations, these traditional risk factors may exclude at-risk groups (i.e. lean NAFLD).

### Fibrosis scores

Of the 16,006 NAFLD cohort at CUIMC, we identified 356 patients with a biopsy-proven NASH by querying pathology reports. We performed fibrosis calculations on the remaining 15,650 patients lacking histology using the FIB-4, APRI, and NAFLD fibrosis score non-invasive tests (NIT)s. We identified 943 patients with scores suggestive of advanced fibrosis, as indicated by an elevated score in ≥2 metrics. 204 patients have advanced fibrosis indicators across all 3 tests and 2,245 patients have a high score in at least 1 metric. Of the patients with advanced fibrosis in ≥2 NITs, 56.5% are diagnosed with DM2 and/or obesity, 46.8% have abnormal liver enzyme levels, 57% are diagnosed with hyperlipidemia, and 78.6% with hypertension (Figure 4). 92.6% of patients with elevated scores in all three calculations have a DM2, obesity, or abnormal liver enzyme diagnosis. The FIB-4 and APRI calculations were most concordant and together identified 778 (82.5%) of patients with elevated NITs in ≥2 tests. Of the discrepancies between the FIB-4 and APRI scores, 151 patients had FIB-4> 3.25 and APRI< 1.0; and 14 had APRI >1.0 and FIB-4< 3.25. We identified 355 patients with a score suggestive of advanced fibrosis using the FIB-4 and NAFLD FS calculators, and 219 patients with high APRI and NAFLD fibrosis scores.

**Figure 4:**
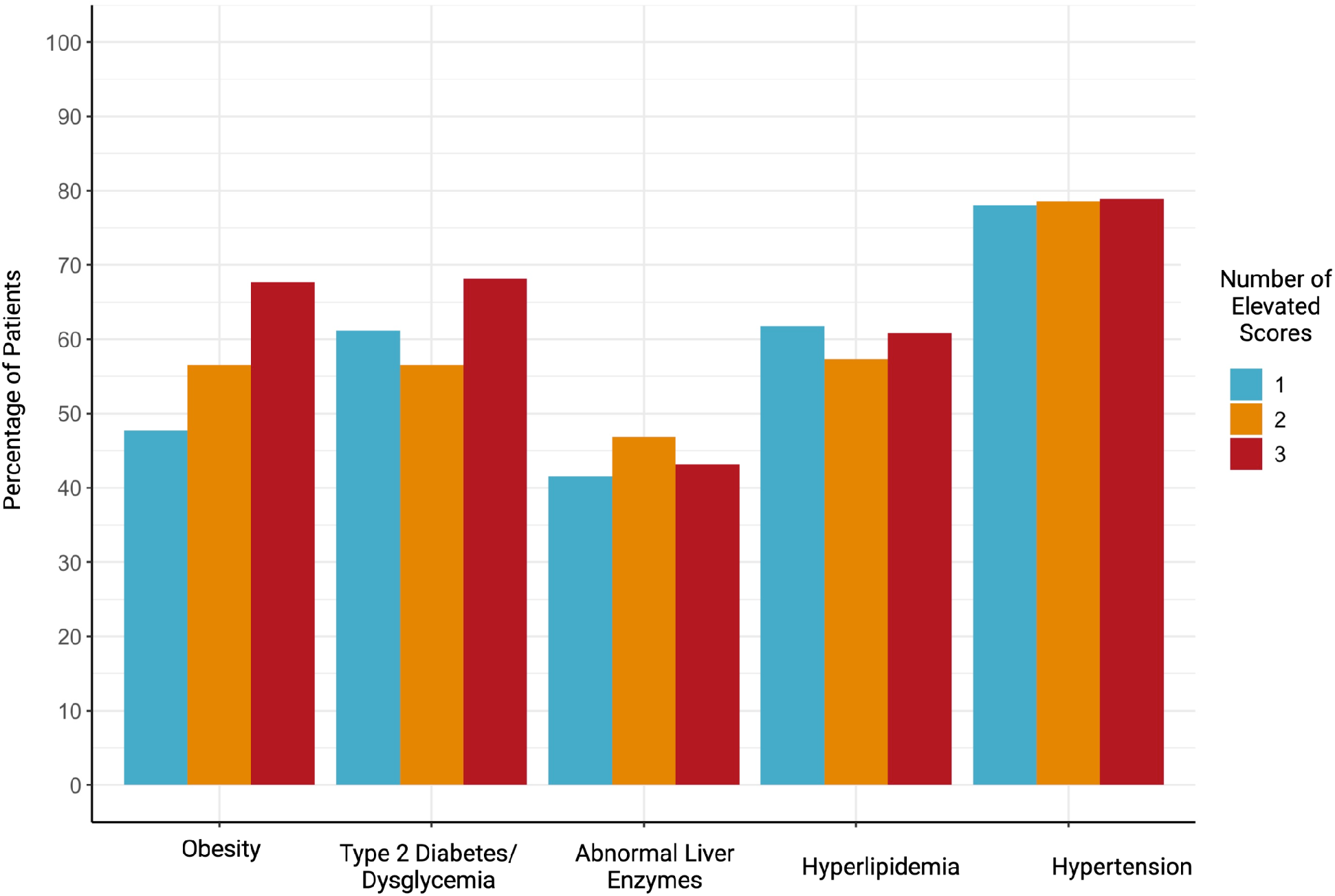
Proportion of patients with select risk indicator diagnoses of those with 1 (n=2,245), 2 (n=942), or 3 (n=204) elevated scores (APRI, NAFLD FS, FIB-4).

### PPV and Sensitivity

Chart review of 200 random patients performed by clinical experts at CUIMC identified 182 individuals correctly discovered by the algorithm as having NAFLD, a PPV of 91%. At the validation sites, our algorithm correctly discovered 15 NAFLD patients (PPV=75%) at UPHS and 17 patients (PPV=85%) at VUMC from a total of 20 patients per site. Algorithmic sensitivity was assessed at CUIMC using clinically-verified NAFLD patients within a registry. When considering the 147 patients with complete data, our NAFLD algorithm attains a sensitivity of 79.6%, identifying 117 patients. The algorithm identifies 147 patients following step 1 (100% sensitivity), 123 after step 2 (83.7% sensitivity), and 117 following step 3 (79.6% sensitivity) (Figure 5). 10 of the 24 patients identified as meeting algorithmic exclusion criteria (step 2) had exclusion codes that could not be clinically-verified during chart review suggesting inconsistencies between the CDW and the clinician-facing system. Our algorithm attains a sensitivity of 85.4% if we drop these 10 patients. When considering all patients within the registry, even those who do not have complete data elements (and are inaccessible to the algorithm), we achieve a sensitivity of 68.3%.

**Figure 5:**
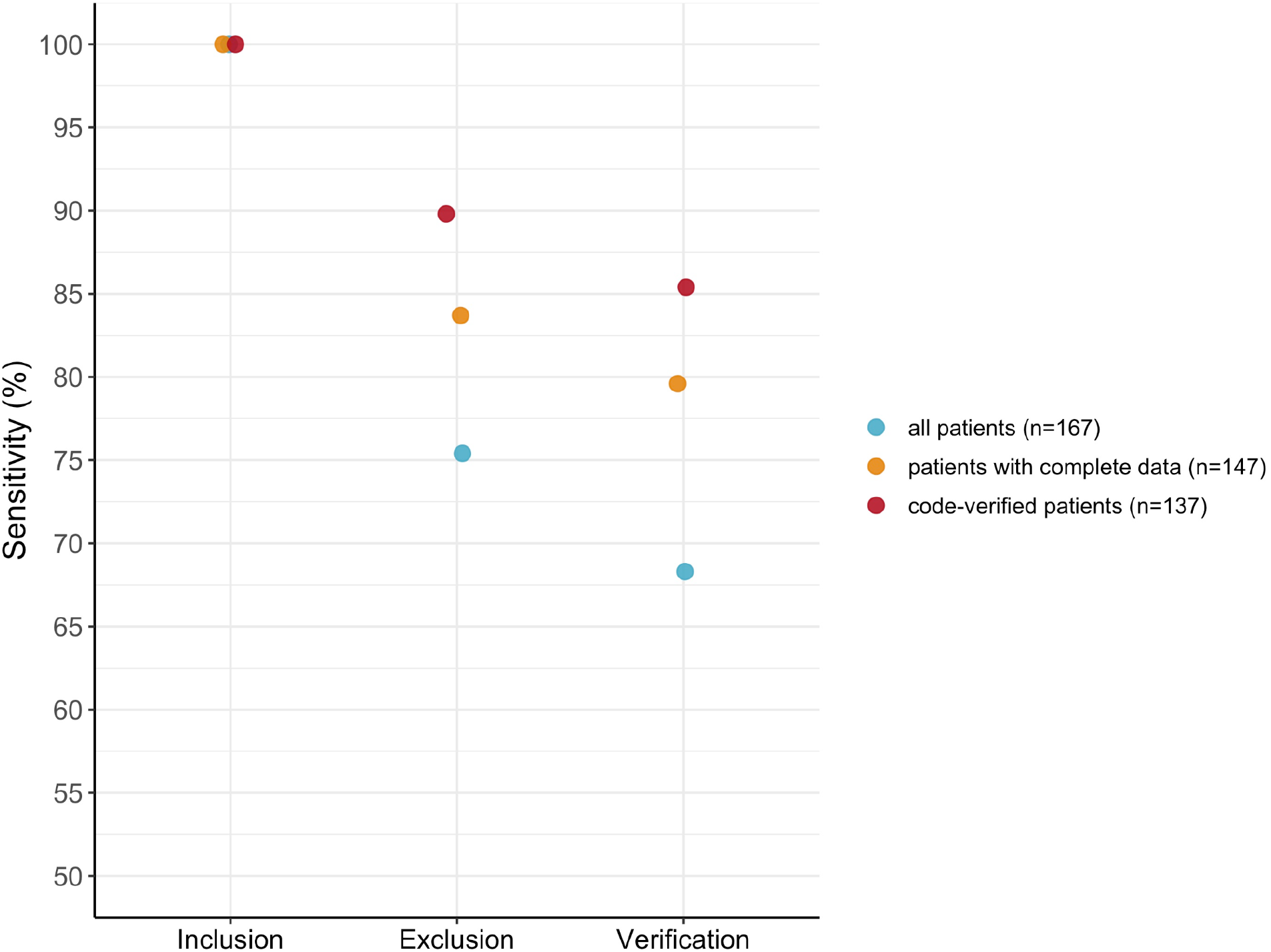
Sensitivity at Columbia University Irving Medical Center (CUIMC) after each stage of the algorithm. Sensitivity was assessed across three categories: 1). using all patients within the NAFLD registry maintained by CUIMC hepatology (blue, n=167), 2). restricting to patients with complete data within the clinical data warehouse (orange, n=147), and 3). restricting to patients with physician-validated exclusion codes (red, n=137). “Inclusion” refers to the identification of NAFLD patients. “Exclusion” is the removal of patients meeting exclusion criteria. “Verification” refers to verification of hepatic steatosis, the final step of our algorithm (see Figure 2).

## Discussion

Identification of patients with under-recognized disease is critical in addressing the NAFLD public health crisis. Reliance on frontline health care recognition may contribute to delays in stratification, monitoring, and referrals for specialty care. While education, awareness campaign,s and resources for provider and patient recognition are necessary, they are not sufficient. Systemic improvements through EHR-based methods can help unburden clinical providers and improve access to advanced therapies. Healthcare systems with interrogatable data provide means of discovering patients at risk for NAFLD and other disease manifestation (^24^). Our work highlights an algorithm that successfully identifies NAFLD and NASH patients across three diverse, urban healthcare systems. 67% of the NAFLD patients discovered at CUIMC were previously undiagnosed by ICD-9/10 codes alone. The algorithm discovers NAFLD patients by first identifying those at risk using diagnoses and risk indicators, then excluding patients with confounding diagnoses, and finally verifying hepatic steatosis. Institutions supporting the OMOP CDM can implement our algorithm to identify at-risk patients for downstream clinical referrals and investigation. Institutions without OMOP can still use the detailed workflow provided with diagnostic codes and search terms for pathology and radiology modalities to assist in patient identification. Given the explosion of biomarkers in the NASH space, we expect future iterations of the algorithm to incorporate additional features.

Our algorithm aggregates large amounts of clinical data and uses imaging or histologic components to verify hepatic steatosis. The algorithm exhibits a high PPV of 91% at CUIMC, 85% at VUMC, and 75% at UPHS showing very good generalizability. The algorithm also incorporates QC measures to reduce the rate of false positives. We found that patients with only one diagnostic code of NAFLD or a risk indicator were predominantly not true NAFLD patients. QC steps requiring a minimum number of unique diagnoses were employed to remove these patients. The algorithm was designed to prioritize PPV so that patients who truly have NAFLD are selected, reducing the false positive rate. This comes with the limitation that not all NAFLD patients will be included, as is reflected by the algorithm’s sensitivity of 79.6%. Our sensitivity analysis highlights circumstances that healthcare centers will need to optimize when applying the algorithm. For example, we identified 10 patients meeting exclusion criteria for whom exclusion codes could not be verified, highlighting a disconnect between data within the CDW and the clinician-facing system. Additionally, missing data elements, particularly radiology and histology reports will affect performance. 12% of NAFLD patients within the CUIMC hepatology registry lacked imaging data within the CDW. Further investigation identified that these patients had imaging performed outside of the healthcare system and are therefore missed by the algorithm. This is particularly significant in referral centers where patients often arrive with outside imaging which is not integrated within the CUIMC CDW. EHRs with centralized, shared data should not have this limitation. Another limitation of the study is small sample sizes for manual chart review at the external validation sites.

Future directions include application to large cohorts like the *All of Us* Research Program and integration of genomic sequencing of groups protected from *and* at-risk for disease progression. Iterations of this algorithm may also be used to identify rare diseases that resemble NAFL, such as familial hyperlipidemias, sub-phenotyping of lean NAFLD for further risk stratification and genomic analysis, and cohort analysis for extrahepatic comorbidities (heart failure, chronic kidney disease). Perhaps most timely, iterative processing of non-invasive scoring systems (FIB-4, NAFLD-FS, APRI) to monitor longitudinal progression in fibrosis will help identify groups for determinants of rapid progression through machine learning methods validated across populations. As circulating and imaging (e.g., elastography) biomarkers for fibrosis become more available, future iterations of this algorithm may identify specific fibrosis stages for clinical trials screening, sub-phenotype analyses, cancer screening, metabolic weight loss and bariatric surgery referrals or liver transplant referrals (Figure 6). As clinical guidelines (^25^) for NAFLD are updated, components may be quickly implemented with algorithmic modification rather than relying on community adoption alone. This may be most advantageous in centers developing innovative linkage to care strategies for diverse populations. Given evolving diagnostic and risk stratification modalities such as serum and imaging based NITs, EHR-based algorithms provide opportunities for discovering patients at highest risk for disease progression and therapeutic intervention for rapid referral and downstream stage-specific intervention.

**Figure 6:**
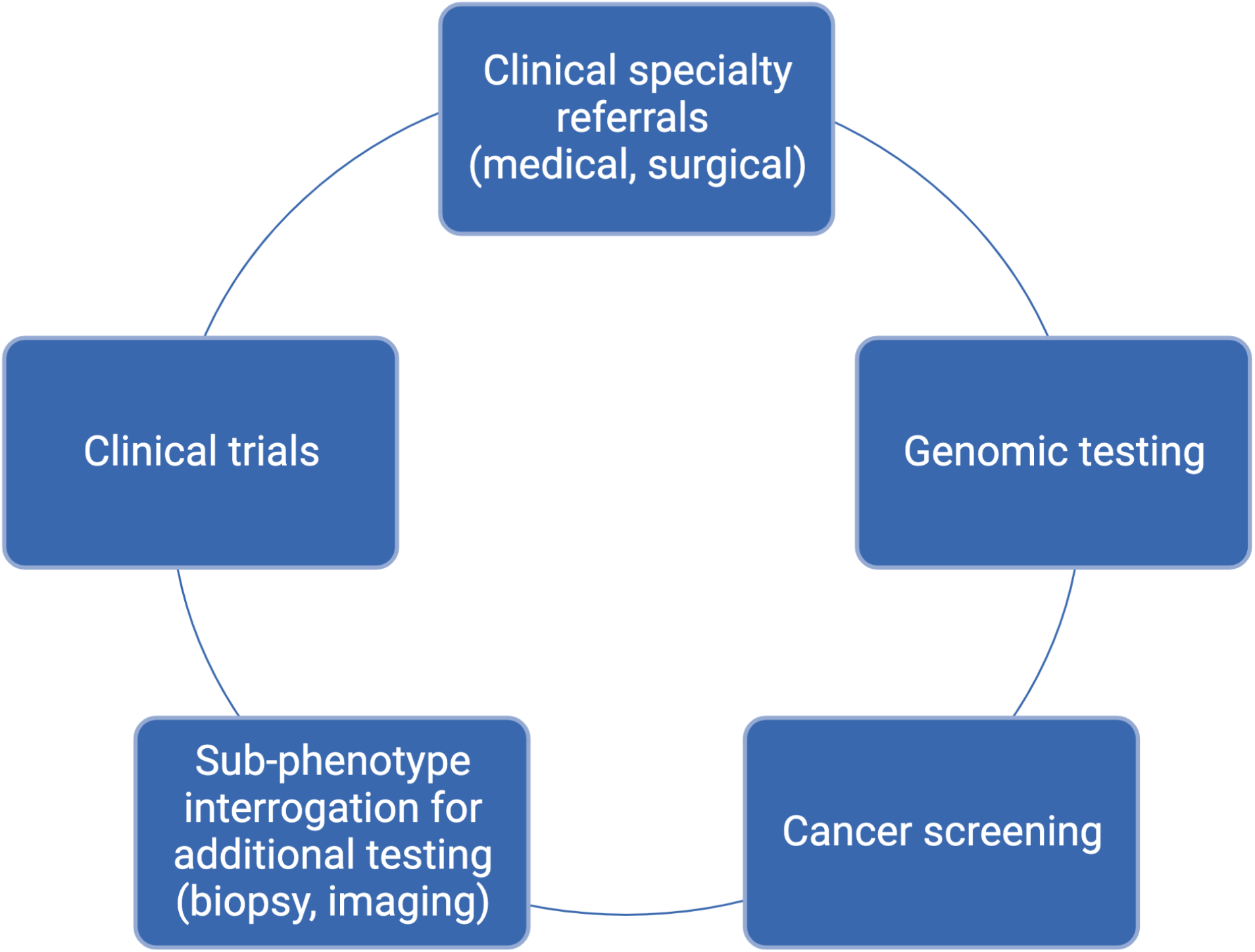
Clinical and translational applications of the NAFLD algorithm.

## Data Availability

Data in the study were extracted from patient electronic health records at Columbia University Irving Medical Center,University of Pennsylvania Healthcare System (UPHS), and Vanderbilt Medical Center (VUMC). These data are not available for public use due to institutional privacy policies and federal regulations.

## Acknowledgements

We would like to thank Joshua C. Denny MD, MS for his scientific contributions to this project as faculty at Vanderbilt University Medical Center prior to joining NIH.

## Abbreviations

EHR: Electronic Health Record
NAFLD: nonalcoholic fatty liver disease
NASH: nonalcoholic steatohepatitis
OMOP: Observational Medical Outcomes Partnership
CDM: Common Data Model
OHDSI: Observational Health Data Sciences and Informatics
A1c: Glycated hemoglobin
T2D: Type 2 Diabetes
DOB: Date of Birth
CUIMC: Columbia University Irving Medical Center
UPHS: University of Pennsylvania Health System
VUMC: Vanderbilt University Medical Center
MRN: Medical Record Number
FIB-4: Fibrosis-4
APRI: Aspartate transaminase to Platelet Ratio Index
NIT: non-invasive test

