## Supplemental Information for "Rapid Identification and Phenotyping of Nonalcoholic Fatty Liver Disease Patients Using an Algorithmic Approach in Diverse, Urban Healthcare Systems"

**Supplemental Digital Content (SDC)**

**Supplementary Equation 1**: FIB-4 score

$$Fib-4=\frac{Age(years)*AST\ Level\ \left(\frac{U}{L}\right)}{Platelet\ Count\ \left(\frac{10^9}{L}\right)*\sqrt{ALT}\left(\frac{U}{L}\right)}$$

**Supplementary Equation 2**: APRI score

$$APRI=\frac{\left(\frac{AST\ Level\frac{IU}{L}}{AST\ (Upper\ Limit\ of\ Normal)\left(\frac{IU}{L}\right)}\right)}{Platelet\ Count\ (10^9L)}*100$$

**Supplementary Equation 3**: NAFLD Fibrosis score

$$NAFLD\ FS=-1.675\ +\ 0.037*\ age(years)+\ 0.094*\ BMI\ \left(\frac{kg}{m2}\right)+\ 1.13*\frac{IFG}{diabetes\ (yes=1,no=2)}+\ 0.99*\frac{AST}{ALT}-\ 0.013*\ platelet\ count\left(\frac{10^9}{L}\right)-\ 0.66*\ albumin(\frac{g}{dL})$$

**Supplementary Table 1: NAFLD risk indicator codes**: This table is divided into sub-tables based on the specific NAFLD risk indicator (**a-e**). Each table lists the OMOP concept name and code id along with the specific diagnostic code and code type. Inclusion criteria for International Classification of Diseases, Ninth and Tenth Revision (ICD-9/10) diagnoses was 1 diagnosis code (dx) occurrence. Any laboratory measures (code type = Logical Observation Identifiers Names and Code (LOINC)) will list the appropriate cutoff for cohort inclusion.

**a. Type 2 diabetes**

| OMOP Concept ID | OMOP Concept Name | Code Type | Specific Code | Criteria for Inclusion |
|---|---|---|---|---|
| 201826 | Type 2 diabetes mellitus | ICD-9/ICD-10 | ICD-9: 250.00, 250.02, ICD-10: E11.00, E11.630 | 1 dx |
| 4193704 | Type 2 diabetes mellitus without complication | ICD-9/ICD-10 | ICD-10: E11.9 | 1 dx |

| | | | | |
|---|---|---|---|---|
| 40482801 | Type II diabetes mellitus uncontrolled | ICD-9/ICD-10 | ICD-9: 250.02 | 1 dx |
| 376065 | Neurologic disorder associated with type 2 diabetes mellitus | ICD-9/ICD-10 | ICD-10: E11.49 | 1 dx |
| 4044391 | Diabetic neuropathy | ICD-9/ICD-10 | ICD-10: E13.40, E08.40 | 1 dx |
| 376979 | Diabetic cataract | ICD-9/ICD-10 | ICD-9: 366.4<br>ICD-10: E08.36 | 1 dx |
| 4009303 | Diabetic ketoacidosis without coma | ICD-9/ICD-10 | ICD-10: E10.10, E13.10, E09.10, E08.10 | 1 dx |
| 4159742 | Diabetic foot ulcer | ICD-9/ICD-10 | ICD-9: E08.621, E13.621 | 1 dx |
| 37018196 | Prediabetes | ICD-9/ICD-10 | ICD-10: R73.03 | 1 dx |
| 192279 | Diabetic renal disease | ICD-9/ICD-10 | ICD-9: 250.4, 249.41, 249.40<br>ICD-10: E13.22, E13.29, E13.21, E09.21, E08.22, E08.21, E13.21, E09.22, E08.29, E09.29 | 1 dx |
| 195771 | Secondary diabetes mellitus | ICD-9/ICD-10 | ICD-9: 249.00, 249.01, 249.80, 249.61, 249.40, 249.90, 249.41, 249.60, 249.40, 249.20, 249.21, 249.81, 249.50, 249.51, 249.70, 249.10, 249.11, 249.91, 249.30, 249.71<br>ICD-10: E08.29, E08.21, E08.22, E08.630, E08.36, E08.618 | 1 dx |
| 201820 | Diabetes mellitus | ICD-9/ICD-10 | ICD-9: 250<br>ICD-10:E13.65, E13.00, 250, E13.649, E08.00, E08.620 | 1 dx |
| 321822 | Peripheral circulatory disorder associated with diabetes mellitus | ICD-9/ICD-10 | ICD-9: 249.70, 250.7, 249.71<br>ICD-10: E13.59, E08.59, E13.51, E08.52, E08.51 | 1 dx |
| 376112 | Diabetic polyneuropathy | ICD-9/ICD-10 | ICD-9: 357.2<br>ICD-10: E13.42, E08.42 | 1 dx |

| | | | | |
|---|---|---|---|---|
| 377552 | Moderate nonproliferative diabetic retinopathy | ICD-9/ICD-10 | ICD-9: 362.05<br>ICD-10: E08.331 | 1 dx |
| 380096 | Proliferative diabetic retinopathy | ICD-9/ICD-10 | ICD-9: 362.02,<br>ICD-10: E08.359, E08.3553, E08.351, E13.359, E13.3592, E13.3593 | 1 dx |
| 380688 | Hypoglycemic coma | ICD-9/ICD-10 | ICD-9: 251.0, 249.31, 249.30 | 1 dx |
| 436940 | Metabolic syndrome X | ICD-9/ICD-10 | ICD-9: 277.7<br>ICD-10: E88.81 | 1 dx |
| 442793 | Diabetic complication | ICD-9/ICD-10 | ICD-9: 250.9, 249.91, 249.80, 249.81, 249.90, 249.81, 249.91<br>ICD-10: E13.8, E08.8, E13.628, E08.69, E13.618, E08.59, E08.638, E08.630, E08.628, E13.69, E13.638, E09.8, E09.69 | 1 dx |
| 443727 | Diabetic ketoacidosis | ICD-9/ICD-10 | ICD-9: 250.1, 249.10, 249.11 | 1 dx |
| 443729 | Peripheral circulatory disorder associated with type 2 diabetes mellitus | ICD-9/ICD-10 | ICD-9: 250.70, 250.6, 250.72, 250.70, 250.6, 249.61, 249.60<br>ICD-10: E11.51, E11.59, E08.49, | 1 dx |
| 443730 | Neurologic disorder associated with diabetes mellitus | ICD-9/ICD-10 | E13.49, E13.49, E08.49, E09.42 | 1 dx |
| 443732 | Disorder due to type 2 diabetes mellitus | ICD-9/ICD-10 | ICD-9:250.90, 250.80, 250.92, 250.82<br>ICD-10: E11.8, E11.69, E11.628, E11.620, E11.638 | 1 dx |
| 443733 | Diabetic oculopathy associated with type 2 diabetes mellitus | ICD-9/ICD-10 | ICD-9: 250.52<br>ICD-10: E11.39, E11.359, E11.319 | 1 dx |
| 443767 | Diabetic oculopathy | ICD-9/ICD-10 | ICD-9: 250.50, 249.51, 250.5, 249.50<br>ICD-10: E13.311, E13.36, E13.39, E13.39, E08.39 | 1 dx |

| | | | | |
|---|---|---|---|---|
| 444369 | Hyperosmolality | ICD-9/ICD-10 | ICD-9: 249.21, 249.20 | 1 dx |
| 4029423 | Hypoglycemic state in diabetes | ICD-9/ICD-10 | ICD-10: E08.65, E13.649, E08.649 | 1 dx |
| 4042728 | Blood glucose abnormal | ICD-9/ICD-10 | ICD-10: R73.09 | 1 dx |
| 4048028 | Diabetic mononeuropathy | ICD-9/ICD-10 | ICD-10: E08.41, E13.41 | 1 dx |
| 4095288 | Diabetic coma with ketoacidosis | ICD-9/ICD-10 | ICD-10: E13.11, E08.11, E09.11 | 1 dx |
| 4096666 | Diabetes mellitus with hyperosmolar coma | ICD-9/ICD-10 | ICD-10: E13.01, E08.01, E13.00 | 1 dx |
| 4114427 | Diabetic neuropathic arthropathy | ICD-9/ICD-10 | ICD-10: E08.618, E13.610, E08.610, E13.618 | 1 dx |
| 4174977 | Diabetic retinopathy | ICD-9/ICD-10 | ICD-9: 362.0, 362.2<br>ICD-10: E13.319, E13.311, E08.311, E08.319, E09.319 | 1 dx |
| 4175440 | Diabetic autonomic neuropathy | ICD-9/ICD-10 | ICD-10: E08.43, E13.43 | 1 dx |
| 4191611 | Diabetic amyotrophy | ICD-9/ICD-10 | ICD-10: E13.44, E08.44 | 1 dx |
| 4214376 | Hyperglycemia | ICD-9/ICD-10 | ICD-10: E11.65, R73.9, E10.65, E13.65 | 1 dx |
| 4226798 | Hypoglycemic coma in diabetes mellitus | ICD-9/ICD-10 | ICD-10: E09.641, E08.641 | 1 dx |
| 4227657 | Diabetic skin ulcer | ICD-9/ICD-10 | ICD-10: E13.622, E08.622 | 1 dx |
| 4308509 | Impaired fasting glycaemia | ICD-9/ICD-10 | ICD-9: 790.21<br>ICD-10: R73.01 | 1 dx |
| 4311629 | Impaired glucose tolerance | ICD-9/ICD-10 | ICD-10: R73.02 | 1 dx |
| 37018196 | Prediabetes | ICD-9/ICD-10 | ICD-10: R73.03 | 1 dx |

| | | | | |
|---|---|---|---|---|
| 3037110 | Hemoglobin A1c/Hemoglobin Total | LOINC | 1558-6 | ≥5.7 |
| 3004410 | Hemoglobin A1c (Glycated) | LOINC | 4548-4 | ≥5.7 |

b. **Obesity**

| **OMOP Concept ID** | **OMOP Concept name** | **Code Type** | **Specific Code** | **Criteria for Inclusion** |
|---|---|---|---|---|
| 3038553 | Body Mass index | LOINC | 39156-5 | >=30 |
| 433736 | Obesity | ICD-9/ICD-10 | ICD-9: 278.00<br>ICD-10: E66.09, E66.8, E66.9 | 1 dx |
| 434005 | Morbid Obesity | ICD-9/ICD-10 | ICD-9: 278.01<br>ICD-10: E66.01 | 1 dx |
| 4060985 | Body mass index 30+ obesity | ICD-9/ICD-10 | ICD-10: V85.38, V85.39, V85.41, Z68.31, Z68.32, Z68.37, Z68.34, Z68.35, Z68.36, Z68.39 | 1 dx |
| 40481140 | Childhood obesity | ICD-9/ICD-10 | ICD-9: V85.54 | 1 dx |
| 4100857 | Extreme obesity with alveolar hypoventilation | ICD-9/ICD-10 | ICD-9: 278.03<br>ICD-10: E66.2 | 1 dx |
| 437525 | Overweight | ICD-9/ICD-10 | ICD-10: E66.3 | 1 dx |
| 4256640 | Body mass index 40+ - severely obese | ICD-9/ICD-10 | ICD-10: Z68.41, V8541 | 1 dx |
| 4097996 | Drug-induced obesity | ICD-9/ICD-10 | ICD-10: E66.1 | 1 dx |

c. **Abnormal Liver Enzymes**

| **OMOP Concept ID** | **OMOP Concept name** | **Code Type** | **Specific Code** | **Criteria for Inclusion** |
|---|---|---|---|---|
| 3006923 | Alanine aminotransferase serum/plasma | LOINC | 1742-6 | >= 40 (2 measurements taken ≥ 6months apart) |

| | | | | |
|---|---|---|---|---|
| 194984 | Disease of Liver | ICD-9/ICD-10 | ICD-9: 573.9, 573.8, 572.8,<br>ICD-10: K76.9, K76.8 | 1 dx |

d. **Hyperlipidemia**

| OMOP Concept ID | OMOP Concept name | Code Type | Specific code | Criteria for Inclusion |
|---|---|---|---|---|
| 3027114 | Cholesterol [Mass/volume] in Serum or Plasma | LOINC | 2093-3 | >200 |
| 3035899 | Cholesterol in LDL [Mass/volume] in Serum or Plasma ultracentrifugate | LOINC | 18261-8 | >=130 |
| 4134862 | Familial hypercholesterolemia | ICD-9/ICD-10 | ICD-10: E78.01 | 1 dx |
| 437827 | Pure hypercholesterolemia | ICD-9/ICD-10 | ICD-9: 272.0<br>ICD-10: E78.00 | 1 dx |
| 432867 | Hyperlipidemia | ICD-9/ICD-10 | ICD-9: 272.4<br>ICD-10: E78.5, E78.4 | 1 dx |
| 438720 | Mixed hyperlipidemia | ICD-9/ICD-10 | ICD-9: 272.2,<br>ICD-10: E78.2 | 1 dx |

e. **Hypertension**

| OMOP Concept ID | OMOP Concept name | Code Type | Specific code | Criteria for Inclusion |
|---|---|---|---|---|
| 320128 | Essential hypertension | ICD-9/ICD-10 | ICD-9: 401.9, 401<br>ICD-10: I10 | 1 dx |
| 312648 | Benign essential hypertension | ICD-9/ICD-10 | ICD-9: 401.1 | 1 dx |
| 4313767 | Chronic peripheral venous hypertension | ICD-9/ICD-10 | ICD-9: 459.30, 459.31, 459.32, 459.33, 459.39 | 1 dx |
| 44782715 | Chronic peripheral venous hypertension with lower extremity complication | ICD-9/ICD-10 | ICD-10: I87.312, I87.393, I87.339, I87.323, I87.329, I87.392, I87.391, I87.399, I87.331, I87.333 | 1 dx |
| 4311246 | Pre-existing hypertension in obstetric context | ICD-9/ICD-10 | ICD-10: O10.013, O10.012, O10.011, O10.019 | 1 dx |

| | | | | |
|---|---|---|---|---|
| 314958 | Benign secondary hypertension | ICD-9/ICD-10 | ICD-9: 405.19, 405.1 | 1 dx |
| 312935 | Venous hypertension | ICD-9/ICD-10 | ICD-10: I87.303, I87.302, I87.309, I87.301 | 1 dx |
| 4064925 | Hypertension screening | ICD-9/ICD-10 | ICD-9: V81.1 | 1 dx |

**Supplementary Table 2: NAFLD diagnosis codes**: This table specifies the ICD-9 or ICD-10 codes for patient inclusion.

| OMOP Concept ID | OMOP Concept Name | Code Type | Specific Code | Criteria for Inclusion |
|---|---|---|---|---|
| 201613 | Chronic nonalcoholic liver disease | ICD-9 | ICD-9: 571.9, 571.8 | 1 dx |
| 40484532 | Nonalcoholic steatohepatitis (NASH) | ICD-9/ICD-10 | ICD-10: K75.81 | 1 dx |
| 4059290 | Steatosis of liver | ICD-9/ICD-10 | ICD-10: K76.0 | 1 dx |
| 194692 | Cirrhosis non-alcoholic | ICD-9/ICD-10 | ICD-9: 571.5 | 1 dx |
| 4064161 | Cirrhosis of liver | ICD-9/ICD-10 | ICD-10: K76.9 | 1 dx |

**Supplementary Table 3: Patient exclusion criteria:** This table is divided into sub-tables based on specific exclusion criteria (**a-f**). Each table lists the OMOP concept name and code id along with the specific diagnostic code and code type. Criteria for cohort exclusion for ICD-9/10 diagnoses was 1 diagnosis (dx).

a. **Alcohol Exclusions**

| OMOP Concept Id | OMOP Concept Name | Code Type | Specific Code | Criteria for Exclusion |
|---|---|---|---|---|
| 433753 | Alcohol abuse | ICD-9/ICD-10 | ICD-9: 305.00<br>ICD-10: F10.10, F10.129, F10.120, F10.19 | 1 dx |
| 435243 | Alcohol dependence | ICD-9/ICD-10 | ICD-9: 303.90<br>ICD-10: F10.20, F10.21, | 1 dx |

| | | | | |
|---|---|---|---|---|
| | | | F10.220, F10.229, F10.231, F10.232, F10.239, F10.24 | |
| 436953 | Continuous chronic alcoholism | ICD-9/ICD-10 | ICD-9: 303.91 | 1 dx |
| 435534 | Nondependent alcohol abuse, continuous | ICD-9/ICD-10 | ICD-9: 305.01 | 1 dx |
| 375519 | Alcohol withdrawal syndrome | ICD-9/ICD-10 | ICD-9: 291.81<br>ICD-10: F10.239, F10.230, F10.232 | 1 dx |
| 196463 | Alcoholic cirrhosis | ICD-9/ICD-10 | ICD-9: 571.2,<br>ICD-10: K70.30, K70.31 | 1 dx |
| 4104431 | Alcohol intoxication | ICD-9/ICD-10 | ICD-9: 303.0<br>ICD-10: F10.120, F10.129, F10.920, F10.929 | 1 dx |
| 433735 | Acute alcoholic intoxication in alcoholism | ICD-9/ICD-10 | ICD-9: 303.00<br>ICD-10: F10.229, F10.220 | 1 dx |
| 441276 | Nondependent alcohol abuse in remission | ICD-9/ICD-10 | ICD-9: 305.03 | 1 dx |
| 201343 | Acute alcoholic liver disease | ICD-9/ICD-10 | ICD-9: 571.1<br>ICD-10: K70.10, K70.11 | 1 dx |
| 439005 | Chronic alcoholism in remission | ICD-9/ICD-10 | ICD-9: 303.93<br>ICD-10: F10.21 | 1 dx |
| 377830 | Alcohol withdrawal delirium | ICD-9/ICD-10 | ICD-9: 291.0<br>ICD-10: F10.231 | 1 dx |
| 437257 | Continuous acute alcoholic intoxication in alcoholism | ICD-9/ICD-10 | ICD-9: 303.01 | 1 dx |
| 376383 | Alcohol-induced organic mental disorder | ICD-9/ICD-10 | ICD-9: 291.8, 291.9<br>ICD-10: F10.288, F10.29, F10.9, F10.94, F10.988, F10.99 | 1 dx |
| 195300 | Alcoholic gastritis | ICD-9/ICD-10 | ICD-9: 535.30, 535.31<br>ICD-10: K29.20, K29.21 | 1 dx |
| 4205002 | Alcohol-induced mood disorder | ICD-9/ICD-10 | ICD-9: 291.89<br>ICD-10: F10.14, F10.24, F10.188, F10.19, F10.288, F10.29, F10.94 | 1 dx |
| 318773 | Dilated cardiomyopathy secondary to alcohol | ICD-9/ICD-10 | ICD-9: 425.5<br>ICD-10: I42.6 | 1 dx |
| 440685 | Nondependent alcohol abuse, episodic | ICD-9/ICD-10 | ICD-9: 305.02 | 1 dx |

| | | | | |
|---|---|---|---|---|
| 193256 | Alcoholic fatty liver | ICD-9/ICD-10 | ICD-9: 571.0<br>ICD-10: K70.0 | 1 dx |
| 201612 | Alcoholic liver damage | ICD-9/ICD-10 | ICD-9: 571.3<br>ICD-10: K70.9 | 1 dx |
| 378726 | Dementia associated with alcoholism | ICD-9/ICD-10 | ICD-9: 291.2<br>ICD-10: F10.27, F10.97 | 1 dx |
| 436585 | Toxic effect of ethyl alcohol | ICD-9/ICD-10 | ICD-9: 980.0<br>ICD-10: T51.0X4A, T51.0X2A, T51.0X1A | 1 dx |
| 40484946 | High alcohol level in blood | ICD-9/ICD-10 | ICD-10: Y90.0, Y90.1, Y90.2, Y90.3, Y90.4, Y90.5, Y90.6, Y90.7, Y90.8 | 1 dx |
| 372607 | Alcohol hallucinosis | ICD-9/ICD-10 | ICD-9: 291.3<br>ICD-10: F10.159, F10.251, F10.951, F10.151 | 1 dx |
| 374623 | Alcohol amnestic disorder | ICD-9/ICD-10 | ICD-9: 291.1<br>ICD-10: F10.96, F10.26 | 1 dx |
| 36714559 | Disorder caused by alcohol | ICD-9/ICD-10 | ICD-10: F10.99, F10.988 | 1 dx |
| 435532 | Episodic chronic alcoholism | ICD-9/ICD-10 | ICD-9: 303.92 | 1 dx |
| 4340383 | Alcoholic hepatitis | ICD-9/ICD-10 | ICD-10: K70.10 | 1 dx |
| 378421 | Alcoholic polyneuropathy | ICD-9/ICD-10 | ICD-9: 357.5<br>ICD-10: G62.1 | 1 dx |
| 435140 | Toxic effect of alcohol | ICD-9/ICD-10 | ICD-9: 980.9, 980.8, 980<br>ICD-10: T51.92XA, T51.8X1A, T51.8X4A, T51.94XA, T51.93XD | 1 dx |
| 46269816 | Ascites due to alcoholic cirrhosis | ICD-9/ICD-10 | ICD-10: K70.31 | 1 dx |
| 441465 | Accidental poisoning by alcoholic beverage | ICD-9/ICD-10 | ICD-9: E860.0 | 1 dx |
| 4042860 | Finding relating to alcohol drinking behavior | SNOMED | 228273003 | 1 dx |
| 433309 | Fetal or neonatal effect of alcohol transmitted | ICD-9/ICD-10 | ICD-9: 760.71 | 1 dx |

| | | | | |
|---|---|---|---|---|
| | via placenta and/or breast milk | | | |
| 4340493 | Alcohol-induced acute pancreatitis | ICD-9/ICD-10 | ICD-10: K85.20, K85.21, K85.22 | 1 dx |
| 441261 | Episodic acute alcoholic intoxication in alcoholism | ICD-9/ICD-10 | ICD-9: 303.02 | 1 dx |
| 4340964 | Alcohol-induced chronic pancreatitis | ICD-9/ICD-10 | ICD-10: K86.0 | 1 dx |
| 442582 | Alcohol-induced psychotic disorder with delusions | ICD-9/ICD-10 | ICD-9: 291.5<br>ICD-10: F10.150, F10.250, F10.950 | 1 dx |
| 436607 | Accidental poisoning by alcohol | ICD-9/ICD-10 | ICD-9: E860.9, E860.8<br>ICD-10: T51.91XA, T51.91XD | 1 dx |
| 4340386 | Alcoholic hepatic failure | ICD-9/ICD-10 | ICD-10: K70.40, K70.41 | 1 dx |
| 435983 | Accidental poisoning with ethyl alcohol | ICD-9/ICD-10 | ICD-9: E860.1<br>ICD-10: T51.0X1A, T51.0X1D | 1 dx |
| 46269835 | Hepatic ascites due to chronic alcoholic hepatitis | ICD-9/ICD-10 | ICD-10: K70.11 | 1 dx |
| 4052945 | Stopped drinking alcohol | SNOMED | 4052946 | 1 dx |
| 440892 | Toxic effect of isopropyl alcohol | ICD-9/ICD-10 | ICD-9: 980.2<br>ICD-10: T51.2X4A, T51.2X2A | 1 dx |
| 4088373 | Alcohol intoxication delirium | ICD-9/ICD-10 | ICD-10: F10.121, F10.221, F10.921 | 1 dx |
| 432609 | Acute alcoholic intoxication in remission, in alcoholism | ICD-9/ICD-10 | ICD-9: 303.03 | 1 dx |
| 4330794 | Alcohol intake exceeds recommended daily limit | ICD-9/ICD-10 | ICD-9: 790.3 | 1 dx |
| 4146660 | Alcohol-induced anxiety disorder | ICD-9/ICD-10 | ICD-10: F10.280, F10.980, F10.180 | 1 dx |
| 45757093 | Alcohol dependence in pregnancy | ICD-9/ICD-10 | ICD-10: O99.310, O99.311, O99.312, O99.313 | 1 dx |

| | | | | |
|---|---|---|---|---|
| 4166129 | Finding of alcohol in blood | ICD-9/ICD-10 | ICD-10: Z02.83, R78.0 | 1 dx |
| 375794 | Alcohol-induced sleep disorder | ICD-9/ICD-10 | ICD-9: 291.82<br>ICD-10: F10.982, F10.282 | 1 dx |
| 4004785 | Fetal alcohol syndrome | ICD-9/ICD-10 | ICD-10: Q86.0 | 1 dx |
| 374317 | Alcohol-induced psychosis | ICD-9/ICD-10 | ICD-10: F10.959, F10.259, F10.159 | 1 dx |
| 440010 | Accidental poisoning by isopropyl alcohol | ICD-9/ICD-10 | ICD-9: E860.3<br>ICD-10: T51.2X1A | 1 dx |
| 1326497 | Alcohol abuse, in remission | ICD-9/ICD-10 | ICD-10: F10.11 | 1 dx |
| 45757783 | Gastric hemorrhage due to alcoholic gastritis | ICD-9/ICD-10 | ICD-10: K29.21 | 1 dx |
| 441761 | Methyl alcohol causing toxic effect | ICD-9/ICD-10 | ICD-9: 980.1 | 1 dx |
| 37016176 | Cerebral degeneration due to alcoholism | ICD-9/ICD-10 | ICD-10: G31.2 | 1 dx |
| 46269818 | Hepatic coma due to alcoholic liver failure | ICD-9/ICD-10 | ICD-10: K70.41 | 1 dx |
| 434217 | Poisoning by alcohol deterrent | ICD-9/ICD-10 | ICD-9: 977.3, E947.3 | 1 dx |
| 4176653 | Alcoholic cerebellar degeneration | ICD-9/ICD-10 | ICD-10: G31.2 | 1 dx |
| 439277 | Alcohol withdrawal hallucinosis | ICD-9/ICD-10 | ICD-10: F10.232 | 1 dx |
| 4078688 | Alcohol myopathy | ICD-9/ICD-10 | ICD-10: G72.1 | 1 dx |
| 4062656 | Alcohol consumption screening | ICD-9/ICD-10 | ICD-10: V79.1 | 1 dx |
| 4005284 | Fetal or neonatal effect of maternal use of alcohol | ICD-9/ICD-10 | ICD-10: P04.3 | 1 dx |
| 436300 | Accidental poisoning by methyl alcohol | ICD-9/ICD-10 | ICD-9: E860.2 | 1 dx |
| 4340385 | Alcoholic fibrosis and sclerosis of liver | ICD-9/ICD-10 | ICD-10: K70.2 | 1 dx |
| 45757131 | Alcohol dependence in childbirth | ICD-9/ICD-10 | ICD-10: O99.314 | 1 dx |

| | | | | |
|---|---|---|---|---|
| 4052946 | Alcohol consumption unknown | SNOMED | 160580001 | 1 dx |
| 4052028 | Alcohol intake within recommended sensible limits | SNOMED | 160593006 | 1 dx |
| 4064179 | Maternal care for (suspected) damage to fetus from alcohol | ICD-9/ICD-10 | ICD-10: O35.4XX0 | 1 dx |
| 4028805 | Alcohol-induced pseudo-Cushing's syndrome | ICD-9/ICD-10 | ICD-10: E24.4 | 1 dx |

b. **Viral Hepatitis Exclusions** Patients who tested with the following criteria for the specific test were excluded from the cohort: Positive, Reactive, Detected, Repeatedly Reactive, Confirmed, Indicated. For tests assessing viral load, patients with values above the baseline for detection were excluded.

| **OMOP concept id** | **OMOP Concept Name** | **Code Type** | **Specific Code** |
|---|---|---|---|
| 3002222 | Hepatitis E virus IgM Ab [Presence] in Serum | LOINC | 14212-5 |
| 3002653 | Hepatitis C virus genotype [Identifier] in Serum or Plasma by Probe and target amplification method | LOINC | 32286-7 |
| 3003867 | Hepatitis E virus IgG Ab [Presence] in Serum | LOINC | 14211-7 |
| 3004347 | Hepatitis D virus Ab [Presence] in Serum | LOINC | 13248-0 |
| 3008075 | Hepatitis C virus RNA [Presence] in Blood by Probe and target amplification method | LOINC | 5010-4 |
| 3013801 | Hepatitis C virus Ab [Presence] in Serum or Plasma by Immunoassay | LOINC | 13955-0 |
| 3014700 | Hepatitis B virus DNA [Units/volume] in Serum | LOINC | 11258-1 |
| 3016770 | Hepatitis C virus RNA [#/volume] (viral load) in Serum or Plasma by Probe and target amplification method | LOINC | 20416-4 |
| 3017143 | Hepatitis C virus Ab [Presence] in Serum | LOINC | 16128-1 |
| 3018447 | Hepatitis C virus RNA [Units/volume] (viral load) in Serum or Plasma by Probe and target amplification method | LOINC | 11011-4 |
| 3018806 | Hepatitis B virus core IgM Ab [Units/volume] in Serum | LOINC | 22319-8 |
| 3019284 | Hepatitis B virus surface Ag [Presence] in Serum | LOINC | 5195-3 |

| | | | |
|---|---|---|---|
| 3019510 | Hepatitis B virus surface Ag [Presence] in Serum or Plasma by Immunoassay | LOINC | 5196-1 |
| 3020316 | Hepatitis A virus IgM Ab [Presence] in Serum or Plasma by Immunoassay | LOINC | 13950-1 |
| 3020978 | Hepatitis B virus genotype [Identifier] in Serum or Plasma by Probe and target amplification method | LOINC | 32366-7 |
| 3021125 | Hepatitis C virus RNA [Presence] in Serum or Plasma by Probe and target amplification method | LOINC | 11259-9 |
| 3022058 | Hepatitis B virus DNA [Presence] in Serum or Plasma by Probe and target amplification method | LOINC | 29610-3 |
| 3022169 | Hepatitis D virus Ab [Units/volume] in Serum by Immunoassay | LOINC | 5200-1 |
| 3022560 | Hepatitis B virus core IgM Ab [Presence] in Serum or Plasma by Immunoassay | LOINC | 24113-3 |
| 3022900 | Hepatitis B virus polymerase DNA [Presence] in Blood by Probe and target amplification method | LOINC | 16934-2 |
| 3023378 | Hepatitis B virus e Ag [Presence] in Serum or Plasma by Immunoassay | LOINC | 13954-3 |
| 3024429 | Hepatitis C virus RNA [Units/volume] (viral load) in Serum or Plasma by Probe with amplification | LOINC | 10676-5 |
| 3025267 | Hepatitis B virus surface Ag [Presence] in Serum or Plasma by Neutralization test | LOINC | 7905-3 |
| 3026432 | Hepatitis C virus RNA [Units/volume] (viral load) in Serum or Plasma by Probe and signal amplification method | LOINC | 29609-5 |
| 3027346 | Hepatitis B virus DNA [#/volume] (viral load) in Serum or Plasma by Probe and target amplification method | LOINC | 29615-2 |
| 3030378 | Hepatitis B virus precore TAG [Presence] in Serum by Probe and target amplification method | LOINC | 33633-9 |
| 3032567 | Hepatitis B virus DNA [Units/volume] (viral load) in Serum or Plasma by Probe and target amplification method | LOINC | 42595-9 |
| 3032823 | Hepatitis C virus RNA [log units/volume] (viral load) in Serum or Plasma by Probe and signal amplification method | LOINC | 42617-1 |
| 3034868 | Hepatitis C virus RNA [log units/volume] (viral load) in Serum or Plasma by Probe and target amplification method | LOINC | 38180-6 |
| 3036806 | Hepatitis B virus e Ab [Presence] in Serum or Plasma by Immunoassay | LOINC | 13953-5 |

| | | | |
|---|---|---|---|
| 3038726 | Hepatitis D virus Ab [Presence] in Serum by Immunoassay | LOINC | 40727-0 |
| 3044784 | Hepatitis B Virus YMDD [Presence] in Serum or Plasma by Probe and target amplification method | LOINC | 43279-9 |
| 3047011 | Hepatitis D virus Ag [Presence] in Serum by Immunoassay | LOINC | 44754-0 |
| 3048505 | Hepatitis B virus DNA [log units/volume] (viral load) in Serum or Plasma by Probe and target amplification method | LOINC | 48398-2 |
| 3049213 | Hepatitis C virus RNA [Presence] in Unspecified specimen by Probe and signal amplification method | LOINC | 48576-3 |
| 3049680 | Hepatitis C virus RNA [Log #/volume] (viral load) in Serum or Plasma by Probe and target amplification method | LOINC | 47252-2 |
| 3052023 | Hepatitis C virus Ab Signal/Cutoff in Serum or Plasma by Immunoassay | LOINC | 48159-8 |
| 3053003 | Hepatitis C virus genotype [Identifier] in Blood by Probe and target amplification method | LOINC | 48574-8 |
| 40757341 | Hepatitis B virus basal core promoter mutation [Identifier] in Serum by Probe and target amplification method | LOINC | 54210-0 |
| 40759633 | Hepatitis E virus IgG Ab [Units/volume] in Serum or Plasma by Immunoassay | LOINC | 56513-5 |
| 40761553 | Hepatitis B virus surface Ag [Units/volume] in Serum | LOINC | 58452-4 |
| 43533679 | Hepatitis C virus NS3 gene mutations detected [Identifier] by Genotype method | LOINC | 73654-6 |
| 43533680 | Hepatitis C virus NS5 gene mutations detected [Identifier] by Genotype method | LOINC | 73655-3 |
| 43534035 | Hepatitis C virus resistance panel by Genotype method | LOINC | 72862-6 |

c. **HIV Exclusion Criteria:** Patients who tested with the following criteria for the specific test were excluded from the cohort: Positive, Reactive, Detected, Repeatedly Reactive, Confirmed, Indicated. For tests assessing viral load, patients with values above the baseline for detection were excluded from the cohort.

| **OMOP Concept Id** | **OMOP Concept Name** | **Code Type** | **Specific Code** |
|---|---|---|---|
| 3000685 | HIV 1 RNA [Presence] in Serum or Plasma by Probe and target amplification method | LOINC | 25835-0 |
| 3004365 | HIV 1 proviral DNA [Presence] in Blood by Probe with amplification | LOINC | 9837-6 |

| | | | |
|---|---|---|---|
| 3010074 | HIV 1 RNA [Log #/volume] (viral load) in Plasma by Probe and signal amplification method | LOINC | 29539-4 |
| 3010747 | HIV 1 RNA [#/volume] (viral load) in Serum or Plasma by Probe and target amplification method | LOINC | 20447-9 |
| 3011325 | HIV 1+2 Ab [Presence] in Serum | LOINC | 7918-6 |
| 3012693 | HIV reverse transcriptase gene mutations detected [Identifier] | LOINC | 30554-0 |
| 3012733 | HIV 2 Ab [Units/volume] in Serum or Plasma by Immunoassay | LOINC | 5224-1 |
| 3013906 | HIV 1 Ab [Presence] in Serum | LOINC | 7917-8 |
| 3014347 | HIV 1 RNA [#/volume] in Serum | LOINC | 21333-0 |
| 3016870 | HIV 1 Ab band pattern [Interpretation] in Serum by Immunoblot | LOINC | 13499-9 |
| 3017675 | HIV 1 Ab [Presence] in Serum or Plasma by Immunoassay | LOINC | 29893-5 |
| 3024449 | HIV 2 Ab [Presence] in Serum or Plasma by Immunoassay | LOINC | 30361-0 |
| 3026532 | HIV 1 RNA [Log #/volume] (viral load) in Plasma by Probe and target amplification method | LOINC | 29541-0 |
| 3031527 | HIV 1 RNA [#/volume] (viral load) in Serum or Plasma by Probe with amplification detection limit = 75 copies/mL | LOINC | 41515-8 |
| 3031839 | HIV 1 RNA [Log #/volume] (viral load) in Serum or Plasma by Probe with amplification detection limit = 1.9 log copies/mL | LOINC | 41516-6 |
| 3032728 | HIV genotype [Susceptibility] in Isolate by Genotype method Narrative | LOINC | 49573-9 |
| 3032965 | HIV 1+2 Ab [Presence] in Unspecified specimen by Rapid immunoassay | LOINC | 49580-4 |
| 3038100 | HIV 1 Ab [Presence] in Serum or Plasma by Immunoblot | LOINC | 5221-7 |
| 3039370 | HIV 2 Ab Signal/Cutoff in Serum or Plasma by Immunoassay | LOINC | 51786-2 |
| 3039421 | HIV 1 RNA [Log #/volume] (viral load) in Serum or Plasma by Probe and target amplification method detection limit = 0.5 log copies/mL | LOINC | 51780-5 |
| 3044830 | HIV protease gene mutations detected [Identifier] | LOINC | 33630-5 |
| 3045827 | HIV phenotype [Susceptibility] | LOINC | 45182-3 |
| 3047064 | HIV 1 proviral DNA [Presence] in Blood by Probe and target amplification method | LOINC | 44871-2 |

| | | | |
|---|---|---|---|
| 3049147 | HIV 1+O+2 Ab [Units/volume] in Serum or Plasma | LOINC | 48346-1 |
| 3053246 | HIV 1+O+2 Ab [Presence] in Serum or Plasma | LOINC | 48345-3 |
| 21494795 | HIV 1 and 2 Ab [Identifier] in Serum, Plasma or Blood by Rapid immunoassay | LOINC | 80203-3 |
| 40760007 | HIV 1+2 Ab+HIV1 p24 Ag [Presence] in Serum or Plasma by Immunoassay | LOINC | 56888-1 |
| 4276586 | Finding of HIV status | ICD-9/ICD-10 | ICD-10: R75 |

d. **Type 1 diabetes exclusions**

| OMOP Concept Id | OMOP Concept Name | Code Type | Specific Codes | Criteria for Exclusion |
|---|---|---|---|---|
| 443412 | Type 1 diabetes mellitus without complication | ICD-9/ICD-10 | ICD-10: E10.9 | 1 dx |
| 4096668 | Type 1 diabetes mellitus with gangrene | ICD-9/ICD-10 | ICD-10: E10.52 | 1 dx |
| 4099214 | Type 1 diabetes mellitus with ulcer | ICD-9/ICD-10 | ICD-10: E10.621,E10.622 | 1 dx |
| 40484648 | Type 1 diabetes mellitus uncontrolled | ICD-9/ICD-10 | ICD-9: 250.03 | 1 dx |
| 201254 | Type 1 diabetes mellitus | ICD-9/ICD-10 | ICD-9: 250.01, 250.03 | 1 dx |
| 201531 | Type 1 diabetes mellitus with hyperosmolar coma | ICD-9/ICD-10 | ICD-9: 250.21 | 1 dx |
| 318712 | Peripheral circulatory disorder associated with type 1 diabetes mellitus | ICD-9/ICD-10 | ICD-9: 250.71, 250.73<br>ICD-10: E10.51, E10.59, E10.52 | 1 dx |
| 373999 | Diabetic oculopathy associated with type 1 diabetes mellitus | ICD-9/ICD-10 | ICD-9: 250.51, 250.53<br>ICD-10: E10.39 | 1 dx |
| 377821 | Neurological disorder associated with type 1 diabetes mellitus | ICD-9/ICD-10 | ICD-9: 250.61, 250.63<br>ICD-10: E10.40, E10.49 | 1 dx |
| 435216 | Disorder due to type 1 diabetes mellitus | ICD-9/ICD-10 | ICD-9: 250.91, 250.81, 250.83, 250.93 | 1 dx |

| | | | | |
|---|---|---|---|---|
| | | | ICD-10: E10.69, E10.8 | |
| 443592 | Hyperosmolality due to uncontrolled type 1 diabetes mellitus | ICD-9/ICD-10 | ICD-9: 250.23 | 1 dx |
| 4063042 | Pre-existing type 1 diabetes mellitus | ICD-9/ICD-10 | ICD-10: O24.03 | 1 dx |
| 4143857 | Amyotrophy due to type 1 diabetes mellitus | ICD-9/ICD-10 | ICD-10: E10.44 | 1 dx |
| 4224254 | Ketoacidotic coma in type 1 diabetes mellitus | ICD-9/ICD-10 | ICD-10: E10.11 | 1 dx |
| 4225055 | Mononeuropathy associated with type 1 diabetes mellitus | ICD-9/ICD-10 | ICD-10: E10.41 | 1 dx |
| 4225656 | Diabetic cataract associated with type 1 diabetes mellitus | ICD-9/ICD-10 | ICD-10: E10.36 | 1 dx |
| 4227210 | Diabetic retinopathy associated with type 1 diabetes mellitus | ICD-9/ICD-10 | ICD-10: E10.319, E10.311 | 1 dx |
| 4152858 | Type 1 diabetes mellitus with arthropathy | ICD-9/ICD-10 | ICD-10: E10.618 | 1 dx |

e. **Other excluding diagnoses**

| OMOP Concept Id | OMOP Concept Name | Code Type | Specific Code | Criteria for Exclusion |
|---|---|---|---|---|
| 192275 | Alpha-1-antitrypsin deficiency | ICD-9/ICD-10 | ICD-9: 273.4, ICD-10: E88.01 | 1 dx |
| 192675 | Biliary cirrhosis | ICD-9/ICD-10 | ICD-9: 571.6 ICD-10: K74.5 | 1 dx |
| 195856 | Cholangitis | ICD-9/ICD-10 | ICD-9: 576.1 ICD-10: K83.0 | 1 dx |
| 4055341 | Calculus of bile duct with cholangitis | ICD-9/ICD-10 | ICD-10: K80.30, K80.34, K80.36, K80.32 | 1 dx |
| 4135822 | Primary biliary cholangitis | ICD-9/ICD-10 | ICD-10: K74.3 | 1 dx |
| 46269831 | Cholangitis due to bile duct calculus with obstruction | ICD-9/ICD-10 | ICD-10: K80.31, K80.33, K80.37, K80.35 | 1 dx |

| | | | | |
|---|---|---|---|---|
| 434614 | Disorder of iron metabolism | ICD-9/ICD-10 | ICD-9: 275.0, 275.09<br>ICD-10: E83.19, E83.10 | 1 dx |
| 436672 | Disorder of copper metabolism | ICD-9/ICD-10 | ICD-9: 275.1<br>ICD-10: E83.00, E83.09 | 1 dx |
| 438721 | Disorder of mineral metabolism | ICD-9/ICD-10 | ICD-9: 275.8, 275.9<br>ICD-10: E83.89, E83.9 | 1 dx |
| 4148231 | Hereditary hemochromatosis | ICD-9/ICD-10 | ICD-9: 275.01<br>ICD-10: E83.110 | 1 dx |
| 4163735 | Hemochromatosis | ICD-9/ICD-10 | ICD-9: 275.02, 275.03<br>ICD-10: E83.111, E83.118, E83.119 | 1 dx |
| 4234997 | Disorder of vein | ICD-9/ICD-10 | ICD-9: 453<br>ICD-10: I87.8, I87.9, | 1 dx |
| 37016193 | Hemochromatosis following repeated red blood cell transfusion | ICD-9/ICD-10 | ICD-10: E83.111 | 1 dx |
| 4031958 | Trace element excess | SNOMED | 238145001 | 1 dx |
| 4043346 | Disorder of thorax | ICD-9/ICD-10 | ICD-10: S23.9XXA, S24.8XXA, S23.29XA, S23.8XXA | 1 dx |
| 4148231 | Hereditary hemochromatosis | ICD-9/ICD-10 | ICD-9: 275.01<br>ICD-10: E83.110 | 1 dx |
| 4064036 | Generalized skin eruption caused by drug and medicament (DRESS syndrome) | ICD-9/ICD-10 | ICD-10: L27.0 | 1 dx |
| 4058694 | Toxic liver disease with cholestasis | ICD-9/ICD-10 | ICD-10: K71.0 | 1 dx |
| 4058695 | Toxic liver disease with fibrosis and cirrhosis of liver | ICD-9/ICD-10 | ICD-10: K71.7 | 1 dx |
| 4316372 | HELLP syndrome | ICD-9/ICD-10 | ICD-10: O14.20, O14.22, O14.24, O14.25, O14.23 | 1 dx |
| 132685 | Severe pre-eclampsia - not delivered | ICD-9/ICD-10 | ICD-9: 642.53 | 1 dx |
| 438490 | Severe pre-eclampsia - delivered | ICD-9/ICD-10 | ICD-9: 642.51 | 1 dx |
| 433536 | Severe pre-eclampsia | ICD-9/ICD-10 | ICD-9: 642.5 | 1 dx |

| | | | | |
|---|---|---|---|---|
| 4057976 | Severe pre-eclampsia with postnatal complication | ICD-9/ICD-10 | ICD-9: 642.54 | 1 dx |
| 439077 | Severe pre-eclampsia - delivered with postnatal complication | ICD-9/ICD-10 | ICD-9: 642.52 | 1 dx |
| 433536 | Severe pre-eclampsia | ICD-9/ICD-10 | ICD-9: 642.50 | 1 dx |
| 4151863 | Congenital abnormality of liver and/or biliary tract | ICD-9/ICD-10 | ICD-10: O26.619 | 1 dx |
| 4062790 | Disease of the digestive system complicating pregnancy, childbirth and/or the puerperium | ICD-9/ICD-10 | ICD-10: O26.613, O99.612, O99.62, O99.63, O99.611, O26.619 | 1 dx |
| 4228429 | Carnitine deficiency | ICD-9/ICD-10 | ICD-10: E71.40 | 1 dx |
| 195223 | Renal carnitine transport defect | ICD-9/ICD-10 | ICD-9: 277.82, 277.81<br>ICD-10: E71.41 | 1 dx |
| 432294 | Iatrogenic carnitine deficiency | ICD-9/ICD-10 | ICD-9: 277.83<br>ICD-10: E71.43 | 1 dx |
| 4261777 | Ruvalcaba-Myhre syndrome | ICD-9/ICD-10 | ICD-9: E71.440 | 1 dx |
| 45773066 | Secondary carnitine deficiency | ICD-9/ICD-10 | ICD-10: E71.448 | 1 dx |
| 45763567 | Carnitine deficiency due to inborn error of metabolism | ICD-9/ICD-10 | ICD-10: E71.42 | 1 dx |
| 436670 | Metabolic disease | ICD-9/ICD-10 | ICD-9: 277.9, 277.89, 277, 277.8,<br>ICD-10: E88.9, | 1 dx |
| 81539 | Mitochondrial cytopathy | ICD-9/ICD-10 | ICD-9: 277.87 | 1 dx |
| 435233 | Disorder of fatty acid metabolism | ICD-9/ICD-10 | ICD-9: 277.85<br>ICD-10: E71.39, E71.318, E71.30 | 1 dx |
| 441268 | Disorder of peroxisomal function | ICD-9/ICD-10 | ICD-9: 277.86<br>ICD-10: E71.548, E71.50 | 1 dx |
| 4079687 | Tumor lysis syndrome | ICD-9/ICD-10 | ICD-9: 277.88<br>ICD-10: E88.3 | 1 dx |
| 4029270 | Carnitine nutritional deficiency | ICD-9/ICD-10 | ICD-9: 277.84 | 1 dx |

| | | | | |
|---|---|---|---|---|
| 444421 | Alagille Syndrome (Congenital malformation syndromes affecting multiple systems) | ICD-9/ICD-10 | ICD-10: Q44.7 | 1 dx |
| 44835070 | Alagille Syndrome (Congenital malformation syndromes affecting multiple systems) | ICD-9/ICD-10 | ICD-9: 759.89 | 1 dx |
| 434615 | Cystic fibrosis | ICD-9/ICD-10 | ICD-9: 277.00 | 1 dx |
| 45576477<br>35207084 | Cystic fibrosis | ICD-9/ICD-10 | ICD-10: E84.9 | 1 dx |
| 435516 | Abetalipoproteinemia, LCAT deficiency | ICD-9/ICD-10 | ICD-9: 272.5<br>ICD-10: E78.6 | 1 dx |
| 134324 | Lipodystrophy | ICD-9/ICD-10 | ICD-9: 272.6<br>ICD-10: E88.1 | 1 dx |
| 375241 | Reye's Syndrome | ICD-9/ICD-10 | ICD-9: 331.81<br>ICD-10: G93.7 | 1 dx |
| 44828573 | Parenteral nutrition | ICD-9/ICD-10 | ICD-10: V58.69 | 1 dx |
| 4082397 | Parenteral nutrition | ICD-9/ICD-10 | ICD-10: Z76.0 | 1 dx |
| 45571391 | Parenteral nutrition | ICD-9/ICD-10 | ICD-10: Z79.891 | 1 dx |
| 45537679 | Parenteral nutrition | ICD-9/ICD-10 | ICD-10: Z79.899 | 1 dx |

f. **Medication Exclusions** Patients prescribed the following medications were excluded from the cohort

| Anti-retroviral Medications | Other Medications |
|---|---|
| atazanavir | Amiodarone |
| darunavir (TMC114) | Tamoxifen |
| fosamprenavir | Methotrexate |
| indinavir | Cytoxan (cyclophosphamide) |
| Lopinavir | Valproate |
| ritonavir | |
| nelfinavir | |
| ritonavir | |

| |
|---|
| saquinavir |
| tipranavir |
| Nucleoside/Nucleotide Reverse Transcriptase Inhibitors (NRTIs) |
| abacavir |
| didanosine (ddI) |
| emtricitabine (FTC) |
| lamivudine (3TC) |
| stavudine (d4T) |
| tenofovir DF |
| zalcitabine (ddC) |
| zidovudine (AZT) |
| Non-Nucleoside Reverse Transcriptase Inhibitors (NNRTIs) |
| delavirdine |
| efavirenz |
| Etravirine |
| nevirapine |
| enfuvirtide (T-20; fusion inhibitor) |
| maraviroc (CCR5 antagonist) |
| raltegravir (integrase inhibitor) |

**Supplementary Table 4: Radiology modalities and key words used to identify hepatic steatosis.**

| Ultrasound | Computerized Tomography (CT) Scan | Magnetic Resonance Imaging (MRI) |
|---|---|---|
| Echogenic (diffusely, increased, heterogeneous) | Hepatic attenuation | Signal intensity |
| Hepatic steatosis | Steatosis | Hepatic steatosis |
| Fatty liver | Fatty change | Nodular |
| Coarsened echotexture | Heterogeneous enhancement | Cirrhotic |
| Nodular | Cirrhosis/cirrhotic | |
| Cirrhotic | Fatty infiltration | |

**Supplementary Table 5: Pathology key words used to identify hepatic steatosis.**

| |
|---|
| Steatosis |
| Steatohepatitis |
| Non-alcoholic steatohepatitis (NASH) |
| Fatty liver |
| Cirrhosis |
| Non-alcoholic fatty liver disease (NAFLD) |

### Extended methods to Manuscript

Algorithm code consists of SQL queries of the structured data and parsing of the unstructured, free text reports. The workflow of the algorithm may be broken down into three main steps, as seen in Figure 2. Step 1 is the inclusion of potential NAFLD patients and is composed of the identification of patients with NAFLD risk indicators (Step 1a) and that of patients with a NAFLD diagnosis (Step 1b). In step 2, non-NAFLD patients meeting select exclusion criteria are removed from the cohort, and hepatic steatosis is verified in Step 3. Each stage of the algorithm flows consecutively so that a patient will not reach step 3 without meeting the criteria of preceding steps. The algorithm was validated in two independent healthcare institutions: University of Pennsylvania Healthcare System (UPHS) in Philadelphia, Pennsylvania, and Vanderbilt University Medical Center (VUMC) in Nashville, Tennessee (see Algorithm Validation). All figures were created using BioRender.com or the ggplot package in R.

#### *Step 1: Identification of NAFLD patients*

Step 1 identifies NAFLD patients and is broken down into 2 sub-steps. In Step 1a, NAFLD patients are identified by the presence of a NAFLD risk indicator, and in Step 1b, by the presence of a NAFLD diagnosis code. All diagnosis codes used in the algorithm and selection criteria for the NAFLD risk indicators are listed in Supplementary Table 1 (S1). The NAFLD diagnosis codes used for patient selection are listed in Supplementary Table 2 (S2). We required patients to be diagnosed with one risk indicator (Supplementary Table 1) or one

NAFLD diagnosis code (Supplementary Table 2) for cohort inclusion, notably inclusive of cirrhosis. NAFLD risk indicators include the following diagnosis categories: type 2 diabetes and dysglycemia (Table S1a), obesity (Table S1b), abnormal liver enzymes (Table S1c), hyperlipidemia (Table S1d), or hypertension (Table S1e). Abnormal liver enzyme diagnosis required patients to have an alanine aminotransferase (ALT) serum/plasma value ≥40 across at least 2 measurements taken at least 6 months apart. Patients with one diagnosis of the specified International Classification of Diseases, Ninth and Tenth Revision, Clinical Modification (referenced as ICD-9/ICD-10 throughout the manuscript) codes were included in the cohort. For laboratory measurements (coded using Logical Observation Identifiers Names and Codes, or LOINC codes), cutoff values for cohort inclusion are listed in the respective tables.

**Step 2: Exclusion of patients with confounding diagnoses**

Following identification of potential NAFLD patients, cases meeting specified exclusion criteria were removed in Step 2 of the algorithm. The exclusion criteria include diagnosis codes for excessive alcohol use, diagnosis of human immunodeficiency virus (HIV), viral hepatitis, type 1 diabetes, or other confounding liver or liver-affecting conditions that may result in secondary hepatic steatosis, including Alpha-1-antitrypsin deficiency, hemochromatosis, and cystic fibrosis. Patients prescribed a hepatotoxic medication associated with steatosis ([1]), such as an anti-retroviral, tamoxifen, or methotrexate, were also excluded. All patient exclusion criteria are listed in Supplementary Table 3. Patients meeting any of the exclusion criteria were removed from our cohort.

**Step 3: Verification of hepatic steatosis**

Radiology and pathology reports, in the form of unstructured or free-text data, from 1980-2016 were used to verify hepatic steatosis in Step 3. Regular expressions, a powerful pattern search language, and tool ([2]), were used in conjunction with specific key terms to identify language and usage context indicative of hepatic steatosis in a string-matching approach. Language for an indicator of NAFLD, as well as of the inflammatory phenotype, NASH, were included. Supplementary Table 4 lists the various radiological modalities and the keywords that were queried in the respective reports. Supplementary Table 5 specifies the key terms used to identify hepatic steatosis from pathology reports obtained via liver biopsy.

**Fibrosis scoring:**

Histologic confirmation is the current standard for verification of NASH. However, biopsies are often underutilized due to their invasive nature. NASH patients were identified from the total pool of patients with verified hepatic steatosis at CUIMC using NASH specific terminology from pathology records. To identify additional patients who may be at risk for fibrotic NAFLD, including NASH, we applied 3 fibrosis scoring metrics on patients lacking histology. These validated metrics include the Fibrosis-4 (FIB-4) ([3]) calculation (Supplementary Equation 1), the aspartate transaminase (AST) to Platelet Ratio Index (APRI) ([4]) calculation (Supplementary equation 2), and the NAFLD Fibrosis score ([5]) (Supplementary equation 3). Data required for these calculations were extracted from patient clinical records. For each required variable, the mean of all measures within 1 year of the date of verified hepatic steatosis was used. For example, given a patient with verified hepatic steatosis on June 20, 2017, the alanine aminotransferase (ALT) value used in the scoring metric was the mean of all available ALT measures from June 20, 2016 to June 20, 2018. R base functions were used to calculate fibrosis scores, and any patient missing the required data elements was excluded from fibrosis scoring. As each of the fibrosis calculations has advantages and disadvantages, we required patients to exhibit a score suggestive of advanced fibrosis using at least 2 of the metrics. Scores indicative of advanced fibrosis are a FIB-4 > 3.25, an APRI >1.0, and a NAFLD FS > 0.675.

The availability of imaging biomarkers (transient elastography, magnetic resonance elastography, etc) as noninvasive estimates of fibrosis were not robustly integrated into the unstructured data at the time of algorithm development for inclusion.

**Chart Review and Algorithmic Performance**

Manual chart review was performed to review data elements used to build the algorithm and to assess algorithmic performance. Random lists of patient Medical Record Numbers (MRNs), identified by the NAFLD algorithm, were used for chart review verification. Provider and admission notes, discharge summaries, endoscopy records, diagnoses, pathology, and radiology reports were all used during chart review.  Over 150 MRNs were reviewed at CUIMC during algorithmic development using both inpatient and outpatient records. This extensive review allowed us to fine-tune criteria for selection of patients within the cohort. Chart review at CUIMC was conducted by two clinical research coordinators, and subsequently verified by a board-certified transplant hepatologist. Chart review at VUMC and UPHS was similarly performed by board-certified transplant hepatologists.

Chart review was also necessary to assess algorithm accuracy. Results of chart review were used to calculate the positive predictive value (PPV) of the algorithm at each of the assessed medical systems. PPV is defined as the proportion of patients identified by the phenotyping algorithm as having the condition, determined by expert chart review. We reviewed the charts of 200 patients, independent of those assessed for diagnostic code selection used to build the algorithm to calculate PPV at CUIMC. Hepatologists at VUMC and UPHS reviewed 20 charts for PPV calculation. Overall, 390 clinical charts were reviewed during algorithm development and validation.
